# Glucagon-Like Peptide-1 Receptor Agonists Across the Heart Failure Spectrum: A Systematic Review and Meta-Analysis

**DOI:** 10.64898/2026.02.10.26345946

**Authors:** Vicky Muller Ferreira, Victor Ayres Muller

**Affiliations:** Independent Researcher, Rio de Janeiro, Brazil

**Keywords:** semaglutide, heart failure with preserved ejection fraction, meta-analysis, cardiovascular outcomes

## Abstract

We performed a systematic review and meta-analysis of randomized controlled trials evaluating glucagon-like peptide-1 receptor agonists (GLP-1 RAs) versus placebo in adults with heart failure (HF), searching PubMed, Cochrane CENTRAL, and ClinicalTrials.gov through February 2026. The primary outcome was the composite of cardiovascular death and first HF hospitalization. Random-effects meta-analysis used restricted maximum likelihood estimation with Hartung-Knapp-Sidik-Jonkman adjustment. We included 14 studies (6 dedicated HF trials and 8 cardiovascular outcomes trial HF subgroup analyses) encompassing 18,558 patients, of whom 2,499 were randomized in dedicated HF trials. The primary composite did not reach statistical significance (hazard ratio [HR] 0.86, 95% confidence interval [CI] 0.73–1.01; P=0.067; I^2^=47%). GLP-1 RAs significantly reduced all-cause mortality (HR 0.87, 95% CI 0.81–0.93; P<0.001; I^2^=0%), major adverse cardiovascular events (HR 0.83, 95% CI 0.73–0.95; P=0.019), and improved Kansas City Cardiomyopathy Questionnaire Clinical Summary Score (+7.4 points, 95% CI 6.3–8.5) and 6-minute walk distance (+17.6 m, 95% CI 13.4–21.7). Excluding the FIGHT trial (acute HFrEF) yielded a significant primary composite (HR 0.83, P=0.011). The mortality signal was driven primarily by CVOT subgroups; the largest dedicated HFpEF trial (SUMMIT) showed numerically higher mortality (HR 1.25). The strongest evidence supports GLP-1 RAs in HFpEF with obesity.

**Highlights:**

- Primary composite of CV death + HHF was not significant (HR 0.86, P=0.067)
- GLP-1 RAs reduced all-cause mortality (HR 0.87) with no heterogeneity
- KCCQ-CSS improved by 7.4 points and 6MWD by 17.6 m in HFpEF trials
- Mortality benefit driven by CVOT subgroups, not dedicated HF trials
- Strongest evidence supports GLP-1 RAs in HFpEF with obesity

## Introduction

Heart failure (HF) remains a major global public health challenge, affecting over 64 million people worldwide and imposing substantial morbidity, mortality, and healthcare costs [1]. Despite advances in neurohormonal blockade for HF with reduced ejection fraction (HFrEF), therapeutic options for HF with preserved ejection fraction (HFpEF) have been historically limited [2,3]. The recognition that obesity and metabolic dysfunction are central to HFpEF pathophysiology has opened new avenues for treatment [4].

Glucagon-like peptide-1 receptor agonists (GLP-1 RAs), initially developed for type 2 diabetes mellitus (T2DM), have demonstrated robust cardiovascular benefits beyond glucose lowering [5]. These agents reduce body weight, improve insulin sensitivity, decrease inflammation, and may exert direct cardioprotective effects through multiple pathways [6,7]. Large cardiovascular outcomes trials (CVOTs) have established the safety and cardiovascular efficacy of several GLP-1 RAs in patients with T2DM [8–12].

Recently, dedicated HF trials have provided pivotal evidence. The STEP-HFpEF and STEP-HFpEF-DM trials demonstrated that semaglutide 2.4 mg significantly improved symptoms, functional capacity, and weight in patients with HFpEF and obesity [13,14]. The SUMMIT trial showed that tirzepatide reduced the composite of cardiovascular death or worsening HF events in HFpEF with obesity [15]. Conversely, the FIGHT trial of liraglutide in acute decompensated HFrEF did not demonstrate clinical benefit [16].

Prior meta-analyses of GLP-1 RAs in HF have been limited by the absence of these landmark trials and by including only a small number of studies [17–19]. With new data from SUMMIT, SELECT HF subgroup, SOUL HF analysis, and FLOW HF subgroup now available, a comprehensive synthesis is warranted.

We therefore conducted a systematic review and meta-analysis to evaluate the efficacy and safety of GLP-1 RAs compared with placebo on cardiovascular outcomes, functional capacity, and patient-reported outcomes in adults with HF across all ejection fraction phenotypes.

## Methods

### Protocol and Registration

This systematic review was prospectively registered with PROSPERO (CRD420261299844) and conducted in accordance with the Preferred Reporting Items for Systematic Reviews and Meta-Analyses (PRISMA) 2020 guidelines [20]. The completed PRISMA checklist is provided in the Supplementary Material.

### Eligibility Criteria

We included randomized controlled trials (RCTs) that enrolled adults (age ≥18 years) with an established diagnosis of HF (any LVEF phenotype: HFrEF ≤40%, HFmrEF 41–49%, or HFpEF ≥50%), randomized to a GLP-1 RA (semaglutide, liraglutide, tirzepatide, dulaglutide, exenatide, lixisenatide, or albiglutide) versus placebo or usual care, with a minimum sample size of 50 participants and follow-up ≥12 weeks, reporting at least one outcome of interest. We included both dedicated HF trials and pre-specified or post-hoc HF subgroup analyses from CVOTs when HF-specific data were separately reported. Exclusion criteria included T1DM as the primary indication, observational studies, single-arm trials, case reports/series, conference abstracts without full publications, and animal or in vitro studies.

### Information Sources and Search Strategy

We searched PubMed (including MEDLINE), Cochrane CENTRAL, and ClinicalTrials.gov from inception through February 3, 2026, without language restrictions. The search strategy combined Medical Subject Headings (MeSH) and free-text terms for GLP-1 RAs (including all individual agent names), heart failure (including phenotype-specific terms), and randomized controlled trials. The full electronic search strategies for all databases are provided in eTable 1 of the Supplementary Material.

### Study Selection

Records were imported, combined, and deduplicated using a multi-pass approach (exact DOI/PMID matching, normalized title matching, and fuzzy matching with manual adjudication). Clearly irrelevant records (animal studies, non-RCTs, non-HF populations) were excluded during initial screening. All 200 records that passed initial filters were independently assessed by both reviewers using the full PICO eligibility framework. Discrepancies were resolved by consensus. Full-text eligibility was assessed for all remaining records.

### Data Extraction

Data were extracted using a standardized form including: study characteristics (design, agent, dose, population, sample size, follow-up duration), participant demographics (age, sex, BMI, LVEF, diabetes status), and outcomes (hazard ratios with 95% confidence intervals for time-to-event outcomes; mean differences with standard deviations or 95% CIs for continuous outcomes; event counts for safety outcomes). For studies reporting only hazard ratios and confidence intervals (without event counts), we computed log(HR) and standard errors using the generic inverse-variance method.

### Risk of Bias Assessment

Risk of bias was assessed using the Cochrane Risk of Bias 2 (RoB 2) tool [21] across five domains: randomization process, deviations from intended interventions, missing outcome data, measurement of the outcome, and selection of the reported result. For CVOT subgroup analyses, an additional consideration was applied for the D5 (selective reporting) domain, as HF outcomes were typically secondary or exploratory endpoints.

### Effect Measures and Statistical Analysis

The primary outcome was the composite of cardiovascular (CV) death and first HF hospitalization. Secondary outcomes included all-cause mortality, MACE (composite of CV death, non-fatal myocardial infarction, and non-fatal stroke), HF hospitalization, CV death, Kansas City Cardiomyopathy Questionnaire Clinical Summary Score (KCCQ-CSS), 6-minute walk distance (6MWD), body weight change, and left ventricular ejection fraction (LVEF) change. Safety outcomes included serious adverse events (SAEs) and treatment discontinuation due to adverse events.

For time-to-event outcomes, we used the generic inverse-variance method with log-transformed hazard ratios and corresponding standard errors. Continuous outcomes were pooled as mean differences. Safety outcomes (count data) used the Mantel-Haenszel method with risk ratios.

Random-effects meta-analysis was performed using restricted maximum likelihood (REML) estimation for the between-study variance (τ^2^) with the Hartung-Knapp-Sidik-Jonkman (HKSJ) adjustment for confidence intervals [22,23]. The HKSJ method provides more reliable confidence interval coverage compared with the DerSimonian-Laird approach, particularly with a small number of studies [24].

Statistical heterogeneity was assessed using I^2^, τ^2^, and Cochran’s Q test, with 95% prediction intervals to characterize the expected range of effects in future settings [25,26].

### Subgroup and Sensitivity Analyses

Pre-specified subgroup analyses examined the influence of HF phenotype (HFpEF vs. HFrEF/mixed), diabetes status, GLP-1 RA agent, and study type (dedicated HF trial vs. CVOT subgroup). Interaction P values were calculated using a meta-regression approach.

Sensitivity analyses included: (1) fixed-effect (common-effect) meta-analysis, (2) exclusion of the FIGHT trial (acute decompensated HFrEF population differing from other studies), (3) exclusion of tirzepatide (dual GIP/GLP-1 receptor agonist), (4) restriction to dedicated HF trials only (excluding CVOT subgroups), and (5) leave-one-out analysis.

### Publication Bias

Publication bias was assessed visually using funnel plots and statistically using Egger’s regression test [27]. The trim-and-fill method was applied to estimate the impact of potential missing studies [28].

### Certainty of Evidence

The certainty of evidence was assessed using the Grading of Recommendations, Assessment, Development, and Evaluations (GRADE) framework [29] across five domains: risk of bias, inconsistency, indirectness, imprecision, and publication bias.

All analyses were performed in R (version 4.4) using the meta [30] and metafor [31] packages. Statistical significance was defined as P < 0.05 (two-sided).

## Results

### Study Selection

The systematic search identified 1,087 records across three databases: PubMed (n=697), Cochrane CENTRAL (n=356), and ClinicalTrials.gov (n=34). After removing 198 duplicates, 889 unique records underwent title and abstract screening. Of these, 689 were excluded, leaving 200 records for full-text assessment. After detailed eligibility evaluation, 186 records were excluded, and 14 studies met all inclusion criteria and were included in the quantitative synthesis (Figure 1).

**Figure 1.**
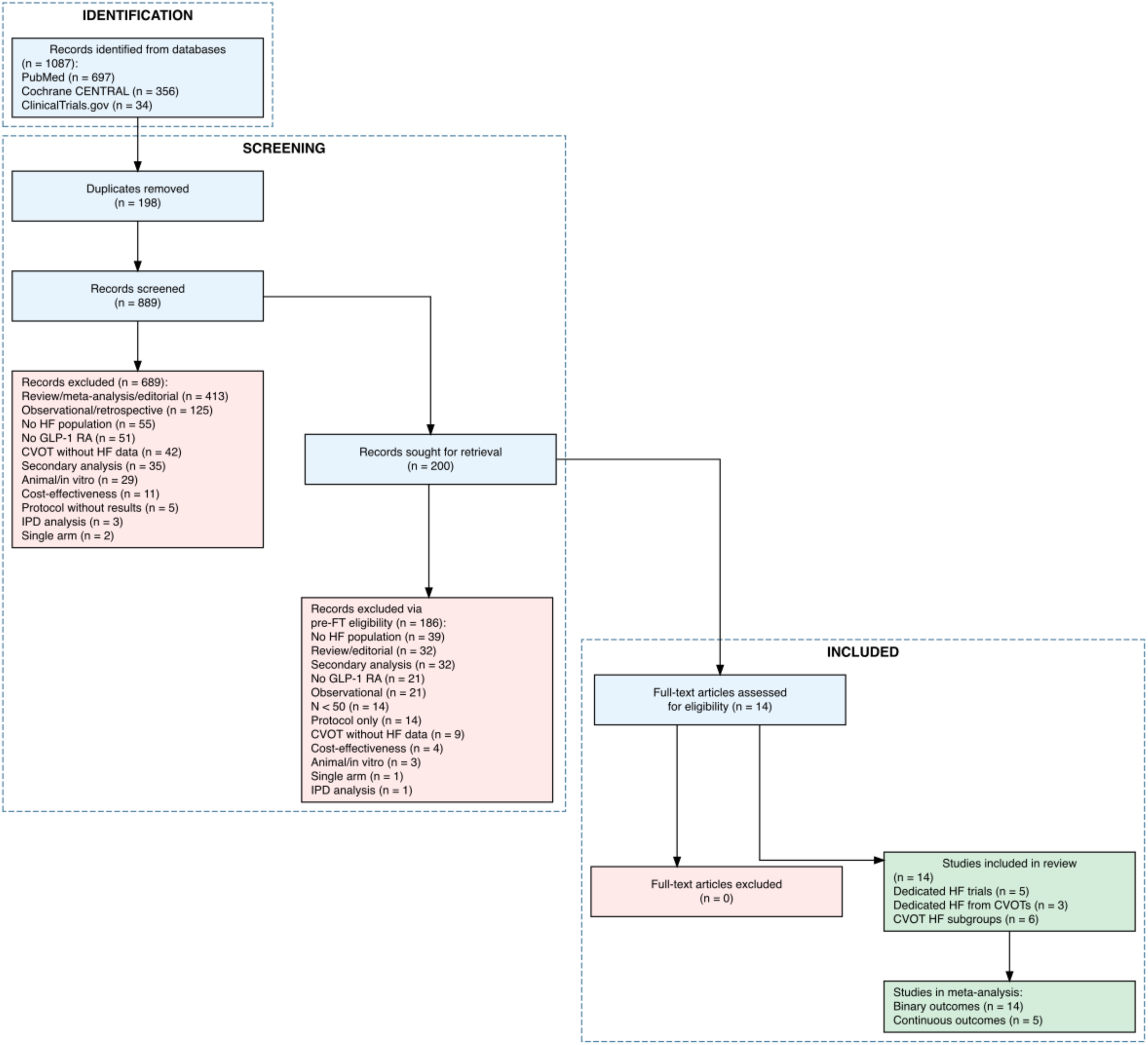
PRISMA 2020 flow diagram of study selection. Figure 1. PRISMA 2020 flow diagram of study selection. A total of 1,087 records were identified across three databases. After deduplication and screening, 14 studies were included in the quantitative synthesis.

### Study Characteristics

The 14 included studies encompassed 18,558 patients in HF subgroups or dedicated HF arms, of whom 2,499 were directly randomized in 6 dedicated HF trials (SUMMIT, STEP-HFpEF, STEP-HFpEF-DM, FIGHT, LIVE, Albiglutide HFrEF), with the remainder enrolled through 8 CVOT HF subgroup analyses (SELECT HF, SOUL HF, FLOW HF, EXSCEL by EF, Harmony Outcomes HF, LEADER HF, REWIND HF, EXSCEL by HF status) (Table 1). For the primary composite, only two studies (SUMMIT and FIGHT; 1,031 patients) provided adjudicated event counts; the remaining six contributed hazard ratios without individual event data. The GLP-1 RAs studied included semaglutide (7 studies), liraglutide (3 studies), exenatide (2 studies), albiglutide (2 studies), dulaglutide (1 study), and tirzepatide (1 study). Mean participant age ranged from 61 to 69 years, and the proportion of women ranged from 15% to 56%. Follow-up ranged from 12 to 273 weeks.

**Table 1.**
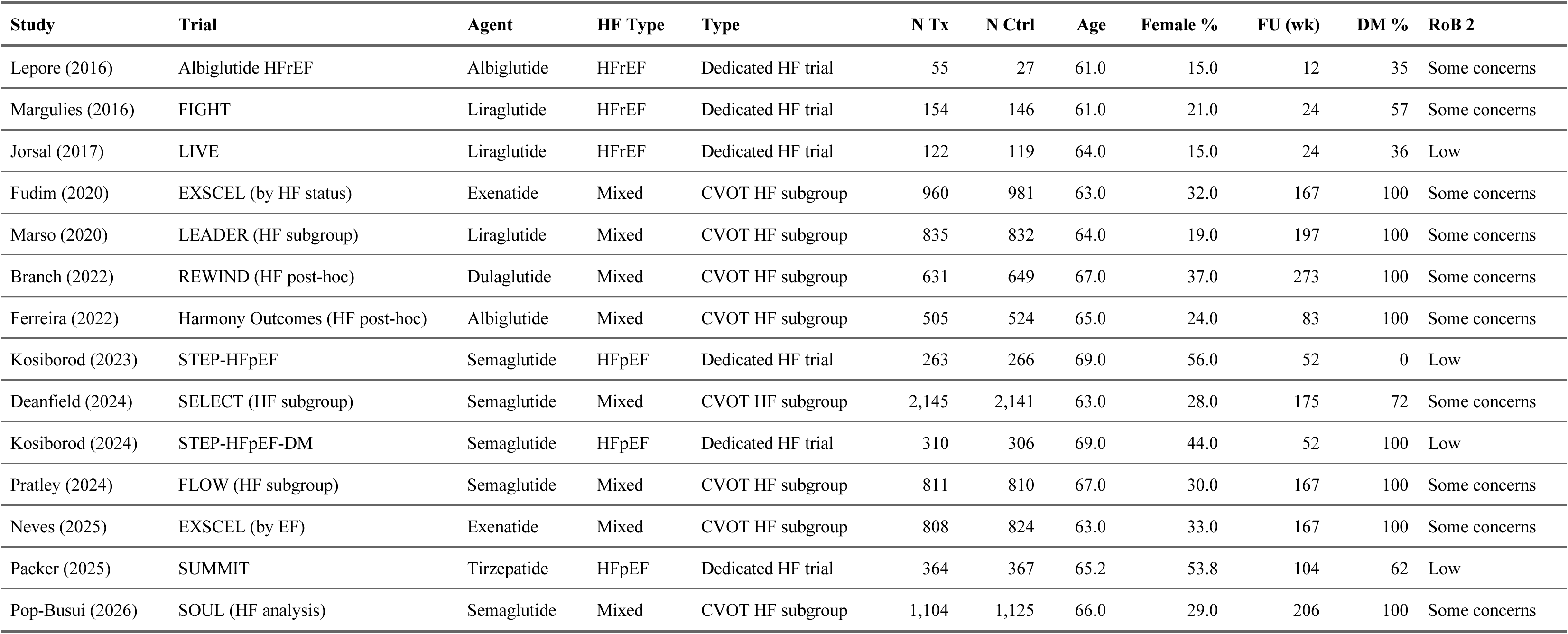
Characteristics of Included Studies.

### Risk of Bias

Four studies were judged as low risk of bias (SUMMIT, STEP-HFpEF, STEP-HFpEF-DM, LIVE), and 10 studies had some concerns, primarily related to post-hoc or subgroup analysis design affecting the selective reporting domain (D5). No studies were rated as high risk of bias. Detailed RoB 2 assessments are presented in eFigures 1–2 and eTable 2.

### Primary Outcome: Composite of CV Death and HF Hospitalization

Eight studies contributed data for the primary composite outcome. The pooled hazard ratio was 0.86 (95% CI 0.73–1.01; P=0.067) using the HKSJ random-effects model, with moderate heterogeneity (I^2^=47%, τ^2^=0.022) (Figure 2, Table 2). The 95% prediction interval ranged from 0.64 to 1.16, suggesting that future studies may show effects ranging from substantial benefit to modest harm.

**Figure 2.**
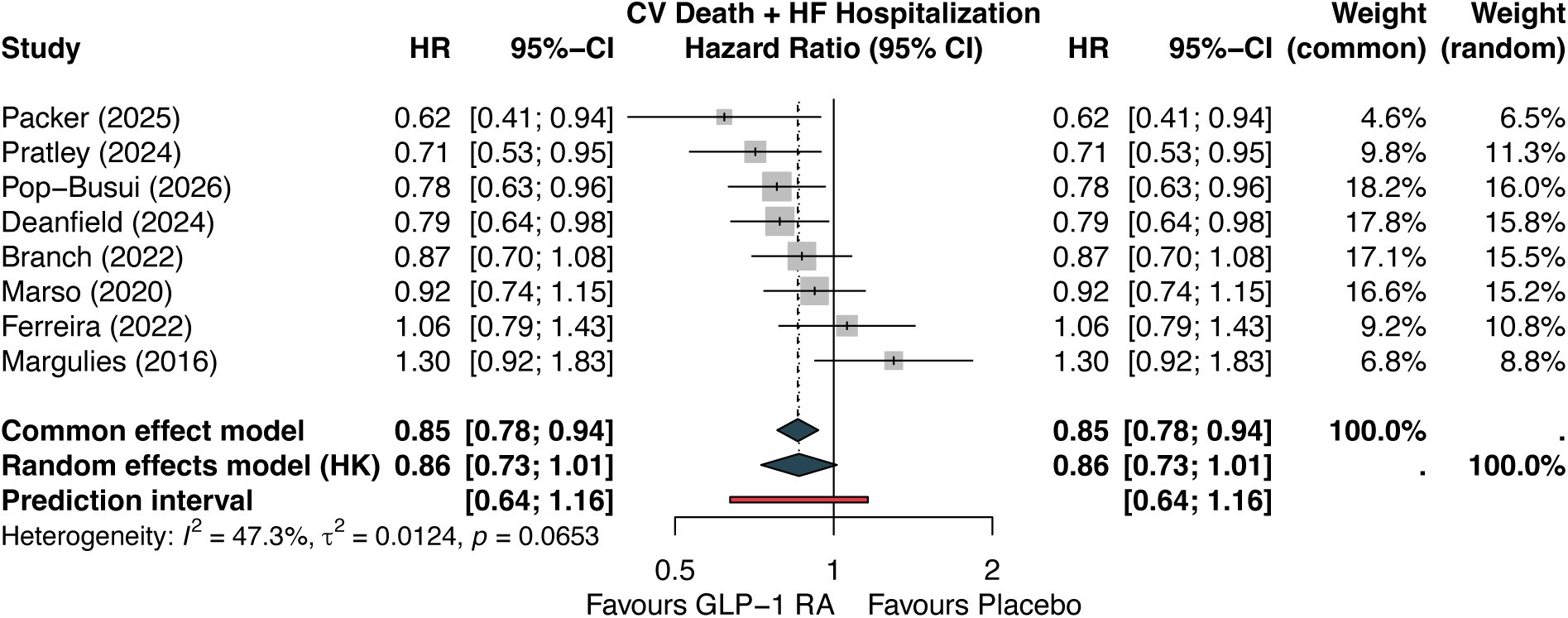
Forest plot — primary composite outcome. Figure 2. Forest plot of the primary composite outcome (cardiovascular death and first heart failure hospitalization). Hazard ratios with 95% confidence intervals from random-effects meta-analysis (REML + HKSJ). The diamond represents the pooled estimate (HR 0.86, 95% CI 0.73–1.01).

**Table 2.**
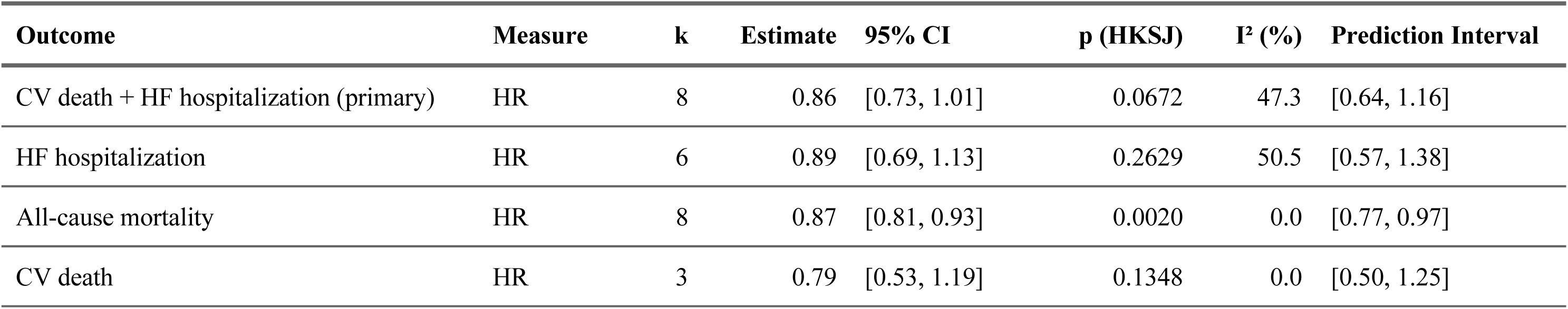
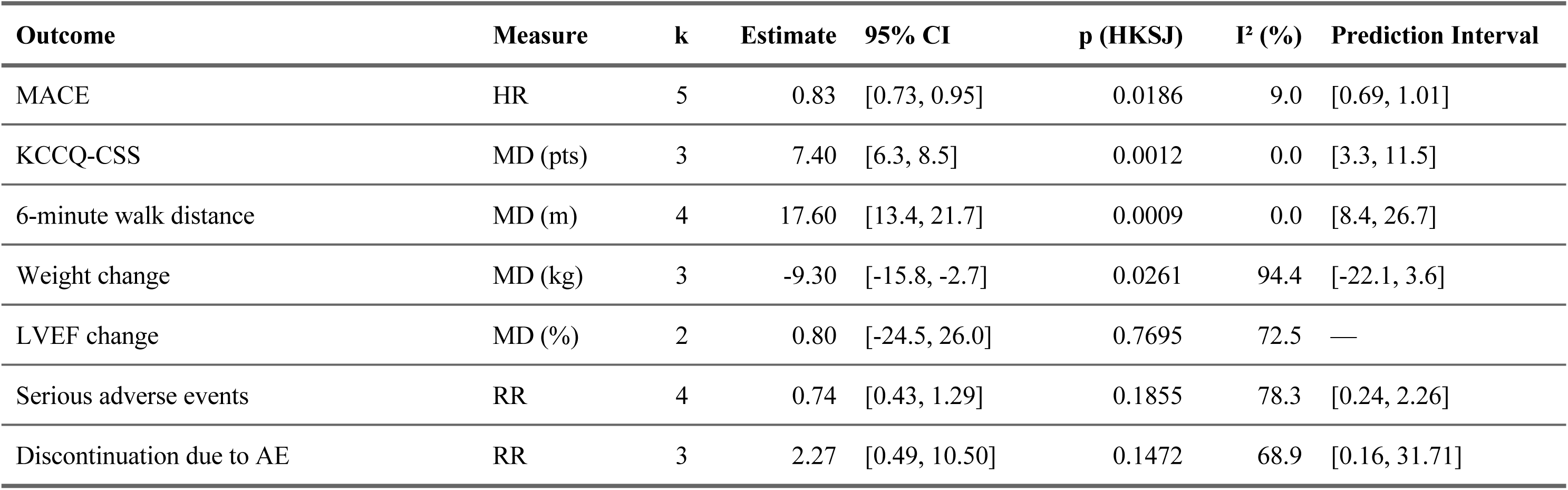
Summary of Meta-Analysis Results.

| Outcome | Measure | k | Estimate | 95% CI | p (HKSJ) | I<sup>2</sup> (%) | Prediction Interval |
| --- | --- | --- | --- | --- | --- | --- | --- |
| CV death + HF hospitalization (primary) | HR | 8 | 0.86 | [0.73, 1.01] | 0.0672 | 47.3 | [0.64, 1.16] |
| HF hospitalization | HR | 6 | 0.89 | [0.69, 1.13] | 0.2629 | 50.5 | [0.57, 1.38] |
| All-cause mortality | HR | 8 | 0.87 | [0.81, 0.93] | 0.0020 | 0.0 | [0.77, 0.97] |
| CV death | HR | 3 | 0.79 | [0.53, 1.19] | 0.1348 | 0.0 | [0.50, 1.25] |
| MACE | HR | 5 | 0.83 | [0.73, 0.95] | 0.0186 | 9.0 | [0.69, 1.01] |
| KCCQ-CSS | MD (pts) | 3 | 7.40 | [6.3, 8.5] | 0.0012 | 0.0 | [3.3, 11.5] |
| 6-minute walk distance | MD (m) | 4 | 17.60 | [13.4, 21.7] | 0.0009 | 0.0 | [8.4, 26.7] |
| Weight change | MD (kg) | 3 | -9.30 | [-15.8, -2.7] | 0.0261 | 94.4 | [-22.1, 3.6] |
| LVEF change | MD (%) | 2 | 0.80 | [-24.5, 26.0] | 0.7695 | 72.5 | — |
| Serious adverse events | RR | 4 | 0.74 | [0.43, 1.29] | 0.1855 | 78.3 | [0.24, 2.26] |
| Discontinuation due to AE | RR | 3 | 2.27 | [0.49, 10.50] | 0.1472 | 68.9 | [0.16, 31.71] |

Individual study estimates ranged from HR 0.62 (SUMMIT) to HR 1.30 (FIGHT), with most studies favoring GLP-1 RAs. Of note, the FIGHT trial enrolled patients with acute decompensated HFrEF and was the only study showing a numerically harmful signal.

### Secondary Outcomes

#### All-Cause Mortality

GLP-1 RAs significantly reduced all-cause mortality across 8 studies (HR 0.87, 95% CI 0.81–0.93; P=0.002; I^2^=0%), with remarkably consistent effects across all studies and a narrow prediction interval of 0.77–0.97 (Figure 3). This corresponds to GRADE moderate certainty evidence (downgraded for indirectness due to CVOT subgroups).

**Figure 3.**
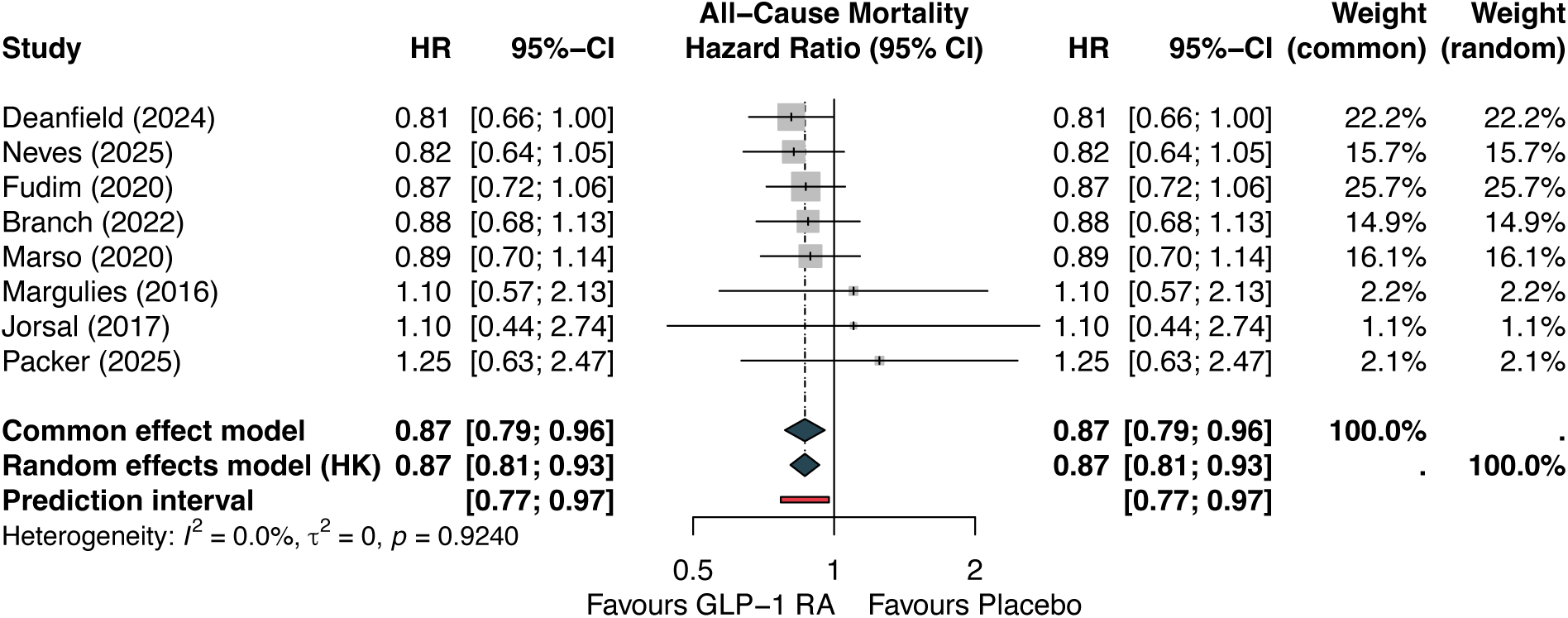
Forest plot — all-cause mortality. Figure 3. Forest plot of all-cause mortality. Pooled HR 0.87 (95% CI 0.81–0.93; I^2^=0%).

#### Major Adverse Cardiovascular Events (MACE)

MACE was significantly reduced in the 5 studies reporting this outcome (HR 0.83, 95% CI 0.73–0.95; P=0.019; I^2^=9%), with consistent effects and GRADE moderate certainty (Figure 4).

**Figure 4.**
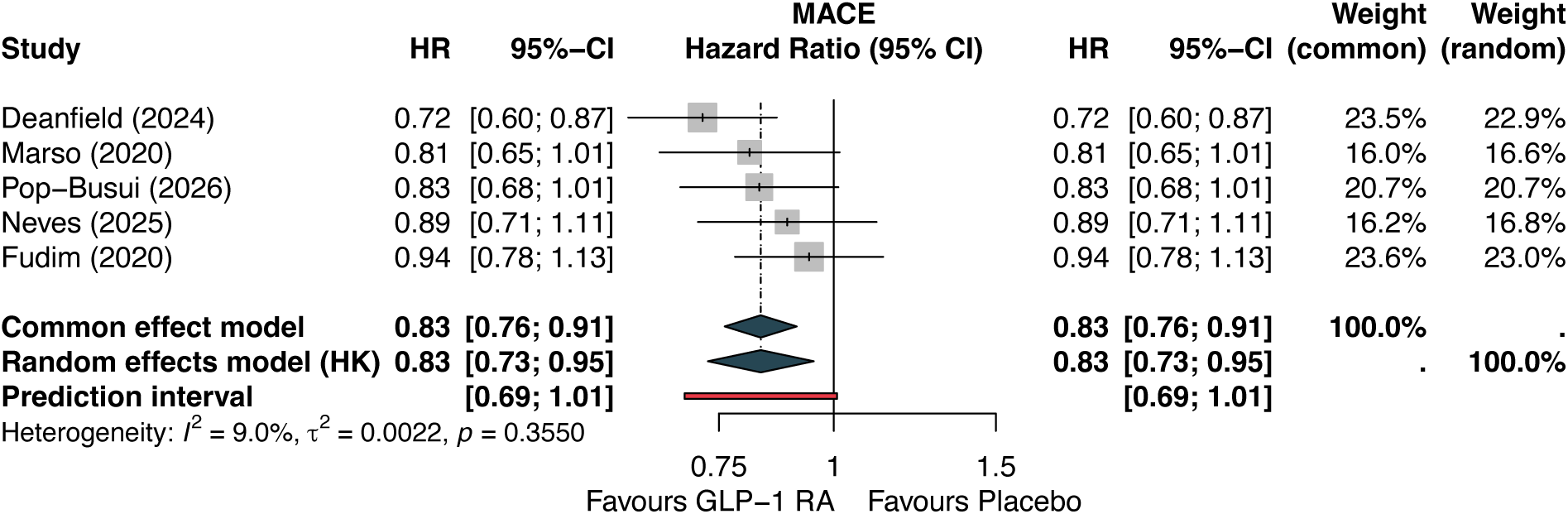
Forest plot — MACE. Figure 4. Forest plot of major adverse cardiovascular events (MACE). Pooled HR 0.83 (95% CI 0.73–0.95; I^2^=9%).

#### HF Hospitalization

HF hospitalization showed a non-significant trend toward reduction across 6 studies (HR 0.89, 95% CI 0.69–1.13; P=0.263; I^2^=51%), with moderate heterogeneity driven by divergent results between dedicated HF trials and CVOT subgroups (Figure 5).

**Figure 5.**
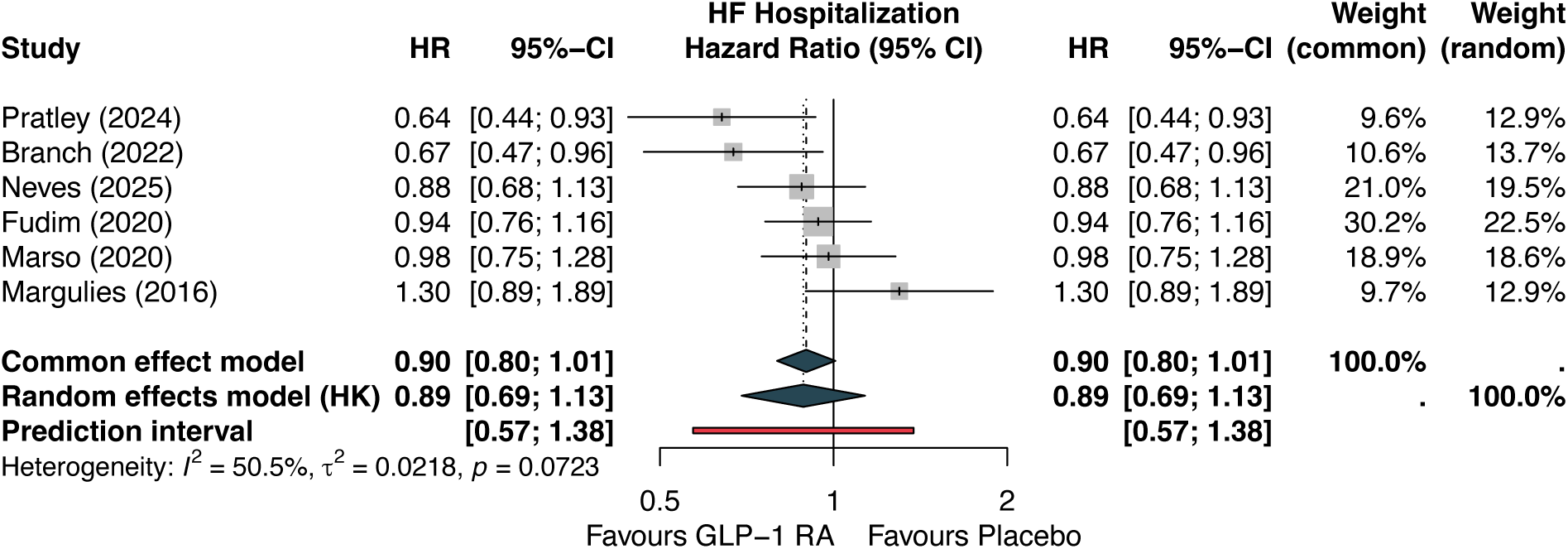
Forest plot — HF hospitalization. Figure 5. Forest plot of heart failure hospitalization. Pooled HR 0.89 (95% CI 0.69–1.13; I^2^=51%).

#### CV Death

CV death showed a non-significant reduction (HR 0.79, 95% CI 0.53–1.19; P=0.135; I^2^=0%; k=3), though limited by the small number of studies contributing to this specific analysis.

#### Functional Outcomes

GLP-1 RAs produced clinically meaningful improvements in both patient-reported outcomes and functional capacity. KCCQ-CSS improved by a mean of 7.4 points (95% CI 6.3–8.5; P=0.001; I^2^=0%; k=3) from the dedicated HFpEF trials (SUMMIT, STEP-HFpEF, STEP-HFpEF-DM), exceeding the minimal clinically important difference of 5 points (Figure 6). The 6-minute walk distance improved by 17.6 m (95% CI 13.4–21.7; P<0.001; I^2^=0%; k=4), also surpassing clinical significance thresholds (Figure 7).

**Figure 6.**
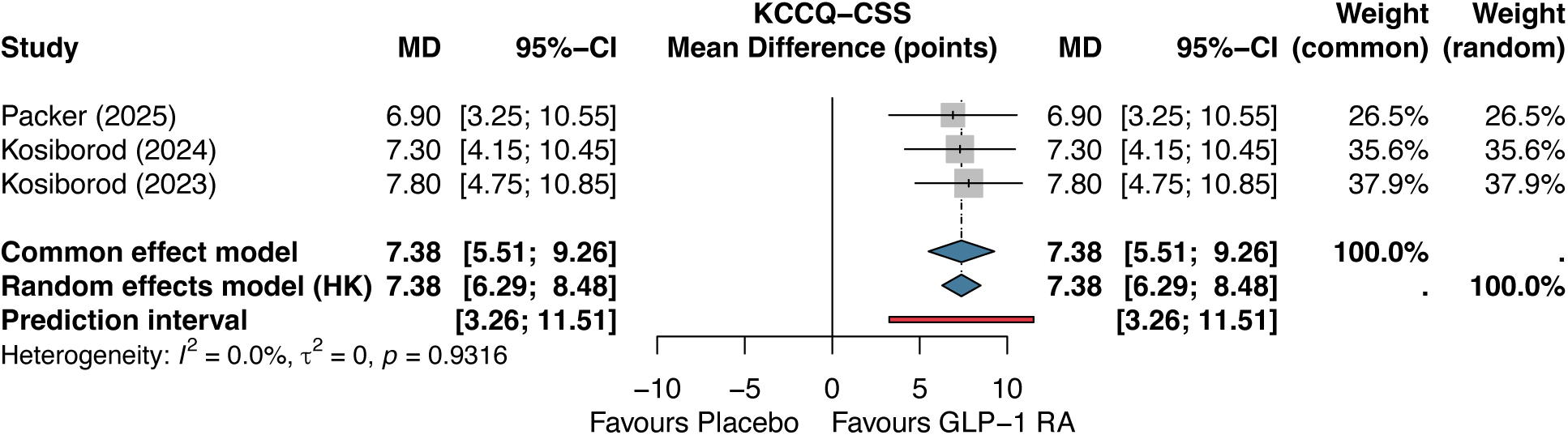
Forest plot — KCCQ-CSS. Figure 6. Forest plot of Kansas City Cardiomyopathy Questionnaire Clinical Summary Score (KCCQ-CSS). Pooled mean difference +7.4 points (95% CI 6.3–8.5; I^2^=0%).

**Figure 7.**
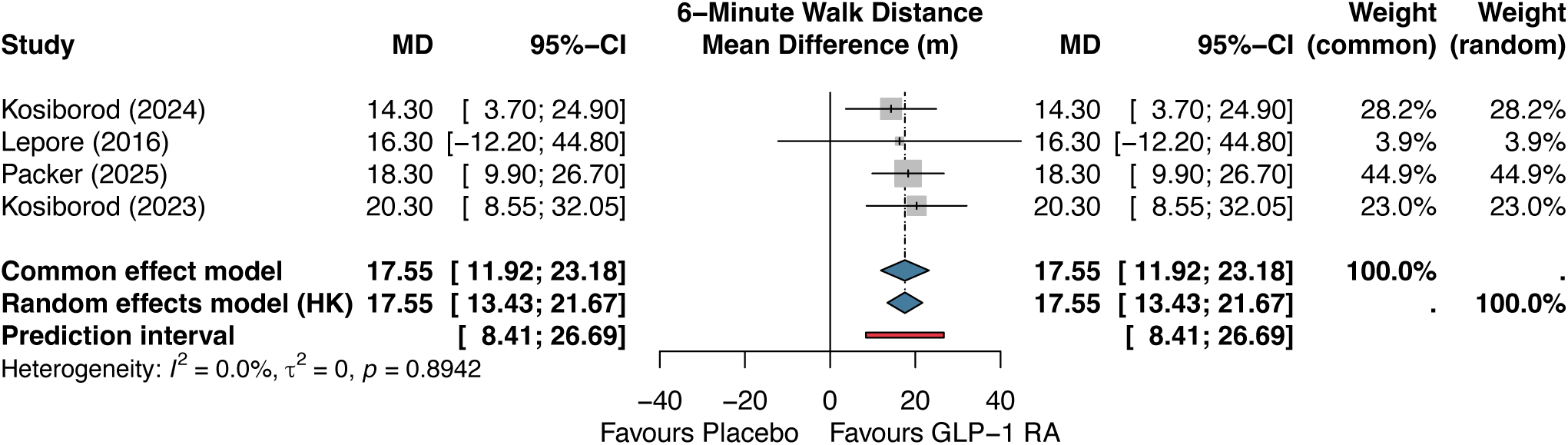
Forest plot — 6MWD. Figure 7. Forest plot of 6-minute walk distance (6MWD). Pooled mean difference +17.6 m (95% CI 13.4–21.7; I^2^=0%).

#### Body Weight and LVEF

Weight reduction was substantial (mean difference –9.3 kg, 95% CI –15.8 to –2.7; P=0.026; I^2^=94%; k=3), though highly heterogeneous, reflecting different agents, doses, and populations. LVEF change was not significantly different between groups (MD 0.8%, 95% CI –24.5 to 26.0; P=0.770; I^2^=73%; k=2).

#### Safety Outcomes

Safety analyses were restricted to dedicated HF trials with available per-arm data. Serious adverse events showed a non-significant trend toward fewer events with GLP-1 RAs (RR 0.74, 95% CI 0.43–1.29; P=0.186; I^2^=78%; k=4), with high heterogeneity reflecting divergent signals between HFpEF trials (favoring GLP-1 RA) and the FIGHT trial in HFrEF (neutral) (Figure 8). Treatment discontinuation due to adverse events was numerically higher with GLP-1 RAs (RR 2.27, 95% CI 0.49–10.50; P=0.147; I^2^=69%; k=3), consistent with known gastrointestinal tolerability concerns.

**Figure 8.**
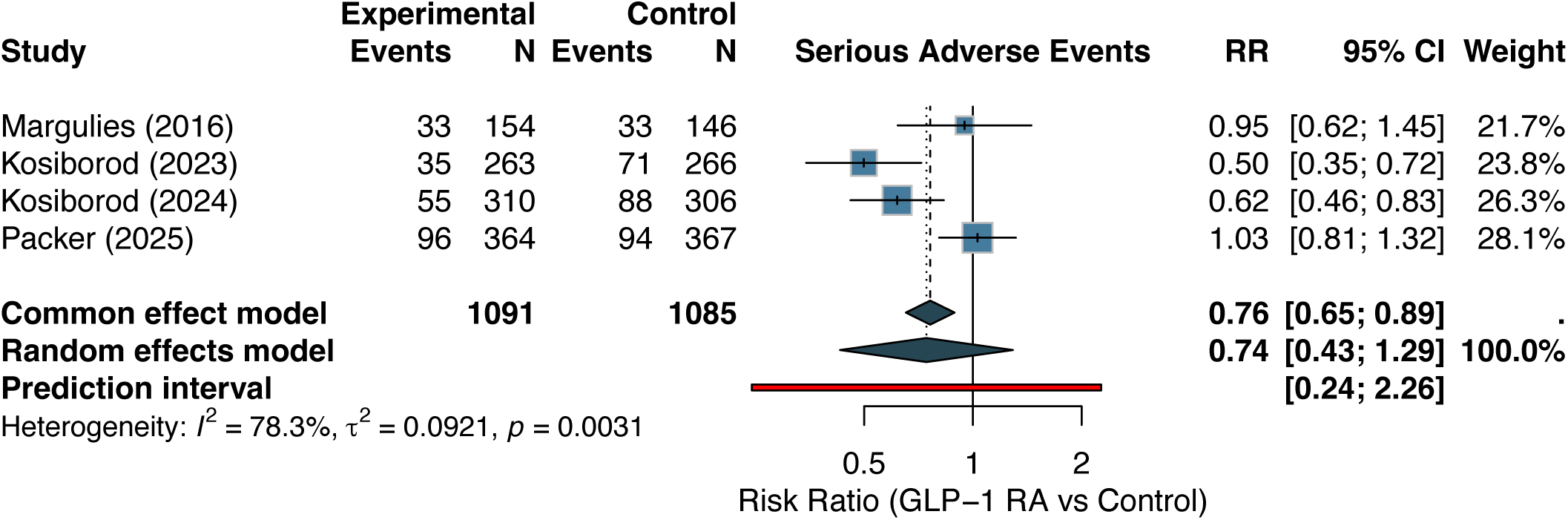
Forest plot — serious adverse events. Figure 8. Forest plot of serious adverse events. Risk ratios from Mantel-Haenszel random-effects meta-analysis. Pooled RR 0.74 (95% CI 0.43–1.29; I^2^=78%).

#### Sensitivity Analyses

Sensitivity analyses for the primary composite outcome are summarized in Table 3. The fixed-effect (common-effect) model yielded a significant pooled HR of 0.85 (95% CI 0.78–0.94; P=0.0006). The difference between fixed- and random-effects results reflects the impact of between-study heterogeneity on confidence interval width under the HKSJ method, which appropriately accounts for this uncertainty with k=8 studies.

**Table 3.**
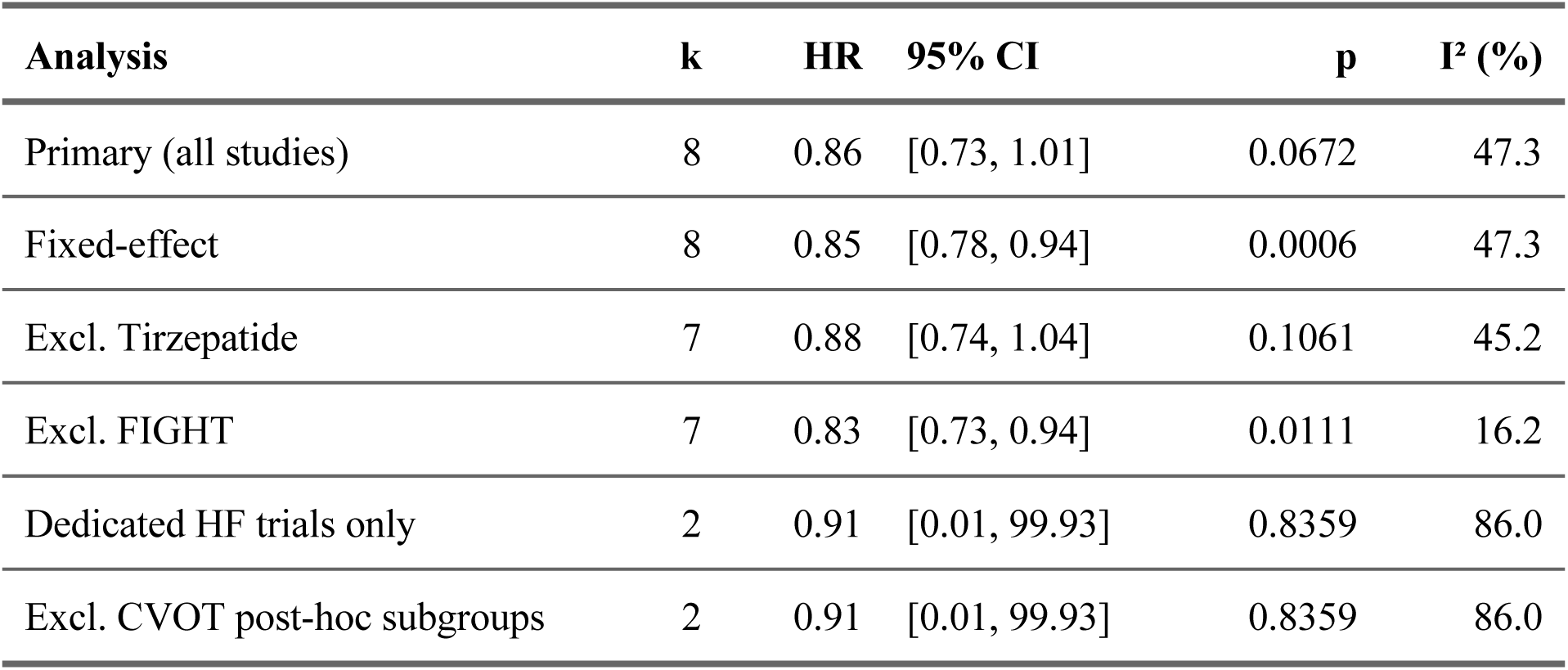
Sensitivity Analyses for the Primary Composite Outcome.

Excluding the FIGHT trial resulted in a significant pooled HR of 0.83 (95% CI 0.73–0.94; P=0.011) with substantially reduced heterogeneity (I^2^=16%), confirming FIGHT as a key source of heterogeneity. Excluding tirzepatide (SUMMIT) had minimal impact (HR 0.88, 95% CI 0.74–1.04; P=0.106). Restricting to dedicated HF trials reporting the primary composite (k=2: SUMMIT and FIGHT) yielded HR 0.84 (95% CI 0.53–1.33; P=0.318; I^2^=66%), with wide confidence intervals reflecting the small number of dedicated trials with primary composite event data and the stark contrast between their results.

Leave-one-out analysis confirmed that no single study disproportionately influenced the overall result beyond FIGHT (eFigure 9).

#### Subgroup Analyses

Subgroup analyses by HF phenotype suggested differential effects: HFpEF-focused studies showed stronger benefit than HFrEF/mixed populations, though formal interaction testing was not statistically significant. Similarly, analyses by GLP-1 RA agent, diabetes status, and study type (dedicated vs. CVOT subgroup) did not reveal significant subgroup interactions, likely reflecting limited statistical power due to the small number of studies within each subgroup (eFigures 10–13).

#### Publication Bias

Visual inspection of the funnel plot for the primary composite outcome showed approximate symmetry (eFigure 7). Egger’s regression test was non-significant (P=0.38), although this test has limited power with fewer than 10 studies (k=8) and these results should be interpreted cautiously [27]. The trim-and-fill analysis estimated no missing studies (eFigure 8).

#### Certainty of Evidence (GRADE)

Using the GRADE framework, the certainty of evidence was moderate for all-cause mortality and MACE, low for the primary composite and HF hospitalization (downgraded for indirectness and imprecision), and moderate for KCCQ-CSS and 6MWD. The primary source of downgrading was indirectness, reflecting the inclusion of CVOT subgroup analyses where HF was not the primary trial endpoint (eTable 3).

## Discussion

This systematic review and meta-analysis provides the most comprehensive synthesis to date of GLP-1 RA effects in heart failure, incorporating 14 studies encompassing 18,558 patients in HF subgroups or dedicated HF arms—of whom 2,499 were directly randomized in 6 dedicated HF trials and 1,031 (in SUMMIT and FIGHT) contributed adjudicated event counts for the primary composite. The primary composite of CV death and HF hospitalization showed a 14% relative risk reduction (HR 0.86) that did not reach statistical significance (P=0.067). Nevertheless, GLP-1 RAs were associated with significant reductions in all-cause mortality (HR 0.87; moderate certainty) and MACE (HR 0.83; moderate certainty), along with clinically meaningful improvements in quality of life (KCCQ-CSS +7.4 points) and functional capacity (6MWD +17.6 m).

The pre-specified HKSJ method provides more accurate confidence interval coverage than the DerSimonian-Laird approach when the number of studies is small [22,23], and the moderate between-study heterogeneity (I^2^=47%) appropriately widened the confidence interval. Notably, the fixed-effect model was significant (P=0.0006), and excluding the FIGHT trial—which enrolled a distinctly different population of acutely decompensated HFrEF patients—yielded significance (P=0.011) with markedly reduced heterogeneity (I^2^=16%).

The FIGHT trial warrants specific discussion. Although FIGHT enrolled acutely decompensated patients, all participants had established chronic HFrEF with acute exacerbation requiring hospitalization, meeting our inclusion criterion of an established HF diagnosis; pre-specified sensitivity analyses excluding this trial were performed to address the resulting clinical heterogeneity. Unlike all other included studies, FIGHT enrolled patients with recent acute HF decompensation and severe HFrEF (mean LVEF 25%), a population with fundamentally different pathophysiology and hemodynamic status [16]. The numerically harmful composite HR of 1.30 in FIGHT contrasts sharply with the consistent benefit seen across all other studies. This divergence may reflect the unsuitability of GLP-1 RA initiation during acute HF decompensation, timing-dependent effects, or the distinct mechanisms underlying acute HFrEF versus chronic HFpEF.

The clinical heterogeneity of the included studies deserves emphasis. This meta-analysis pools data from populations as disparate as obese patients with chronic HFpEF (SUMMIT, STEP-HFpEF), acutely decompensated HFrEF patients (FIGHT), and diabetes-focused CVOTs where HF was a secondary classification (LEADER, EXSCEL). While precedent exists for class-level pooling in HF meta-analyses—SGLT2 inhibitor meta-analyses similarly combined dedicated HF trials with CVOT HF subgroups—an important distinction is that the SGLT2i evidence base included 5 large dedicated HF trials (DAPA-HF, EMPEROR-Reduced, DELIVER, EMPEROR-Preserved, SOLOIST-WHF) with standardized composite endpoints and adjudicated events, whereas only 2 of our 8 studies reporting the primary composite (SUMMIT and FIGHT) provide event counts from dedicated HF populations. The 95% prediction interval of 0.64–1.16 is perhaps the most honest summary of the expected treatment effect range, indicating that while most future HF studies of GLP-1 RAs would likely show benefit, a study in an unfavorable population (e.g., acute HFrEF) could plausibly show harm.

The signal for benefit appears strongest in HFpEF with concurrent obesity, supported by the dedicated trials. The SUMMIT trial demonstrated a striking 38% reduction in the composite of CV death or worsening HF with tirzepatide [15], while the STEP-HFpEF trials showed substantial improvements in symptoms and functional capacity with semaglutide [13,14]. This is mechanistically plausible: obesity-related HFpEF is characterized by systemic inflammation, metabolic dysfunction, increased epicardial fat, and volume overload—all targets of GLP-1 RA therapy [4].

An important finding is the Harmony Outcomes HF subgroup analysis, which showed an HR of 1.06 (95% CI 0.79–1.43) for the composite in patients with established HF [32], contrasting with significant cardiovascular benefit in the overall CVOT population. This suggests that the cardiovascular benefits of albiglutide in the broader T2DM population may not extend to those with established HF, or alternatively, that the post-hoc subgroup analysis was underpowered to detect a modest effect. Notably, several major CVOTs—including ELIXA (lixisenatide) [33], SUSTAIN-6 (semaglutide) [34], and PIONEER 6 (oral semaglutide) [35]—were not included because HF-specific subgroup data were not separately published; their inclusion might have strengthened or attenuated the pooled effects.

Semaglutide was the most studied agent, contributing to 7 of 14 included studies across both dedicated HF trials and CVOT subgroups. The consistent benefit observed across these studies—spanning different doses (1.0 mg, 2.4 mg, and oral), populations (HFpEF, mixed), and trial designs—strengthens the evidence for a class effect, with semaglutide having the most robust evidence base. Tirzepatide, as a dual GIP/GLP-1 receptor agonist, showed the largest effect in SUMMIT, though the contribution of GIP receptor agonism to HF outcomes remains to be fully elucidated.

The all-cause mortality finding (HR 0.87; I^2^=0%) warrants nuanced interpretation despite its statistical consistency. The zero heterogeneity reflects the fact that all individual study point estimates fell within a narrow range—but this pooled signal was driven primarily by CVOT subgroup analyses, where the mortality benefit may reflect the broader cardiovascular effects of GLP-1 RAs in patients with diabetes and established CVD rather than HF-specific mechanisms. Importantly, among dedicated HF trials, SUMMIT showed numerically higher all-cause mortality with tirzepatide (HR 1.25; 19 vs. 15 deaths), and both FIGHT (HR 1.10) and LIVE (HR 1.10) showed similar non-significant trends toward harm. The pooled mortality benefit should therefore be interpreted cautiously: it remains uncertain whether GLP-1 RAs reduce mortality through direct HF mechanisms or through broader cardiometabolic pathways captured in CVOT populations. The GRADE certainty was rated moderate (downgraded once for indirectness due to CVOT subgroups).

Several limitations should be acknowledged. First, CVOT subgroup analyses introduce indirectness, as HF was not the primary trial endpoint and subgroup analyses may have limited power and post-hoc design. Second, individual patient data were not available, precluding exploration of patient-level predictors of response and introducing the risk of ecological fallacy in subgroup analyses. Third, trial sequential analysis was not feasible for the primary composite because most studies reported hazard ratios without event counts. Fourth, safety data were limited to dedicated HF trials, as CVOTs did not report safety endpoints for HF subgroups. Fifth, the search was restricted to published studies indexed in the searched databases. Sixth, the short follow-up of some trials (12–52 weeks) may not capture longer-term effects. Seventh, two EXSCEL subgroup analyses were included (Neves et al., stratified by LVEF; Fudim et al., stratified by HF status), which derive from the same parent trial with partially overlapping populations; this may introduce double-counting for secondary outcomes (all-cause mortality, MACE, HF hospitalization) to which both contribute, though neither contributes to the primary composite. Finally, the substantial weight reduction observed (−9.3 kg) makes it difficult to disentangle the direct cardiac effects of GLP-1 RAs from the indirect benefits of weight loss.

### Clinical Implications

These findings support the emerging role of GLP-1 RAs in the management of HF, particularly in patients with HFpEF and obesity—a population with limited therapeutic options. The 2023 ESC focused update already acknowledged the potential of semaglutide in obese HFpEF patients [36], and our meta-analysis provides quantitative evidence to support this positioning. For HFrEF, the evidence remains inconclusive and suggests caution, particularly in the acute decompensated setting. The significant all-cause mortality reduction is a compelling finding that, if confirmed in future dedicated HF mortality trials, could fundamentally expand the treatment paradigm for HF.

Future research should prioritize dedicated HF mortality trials with GLP-1 RAs, head-to-head comparisons with SGLT2 inhibitors (whose HF benefits are established [37–40]), combination studies, and longer-term safety data. The ongoing STRIDE-HF and SELECT-HF Phase 3 trials will provide additional evidence.

## Conclusions

In this systematic review and meta-analysis of 14 studies encompassing 18,558 patients across 6 dedicated HF trials and 8 CVOT HF subgroup analyses, the primary composite of CV death and HF hospitalization did not reach statistical significance (HR 0.86, 95% CI 0.73–1.01; P=0.067). GLP-1 RAs were associated with significant reductions in all-cause mortality (HR 0.87, I^2^=0%) and MACE (HR 0.83), though the mortality benefit was driven primarily by CVOT subgroups rather than dedicated HF trials; the largest dedicated HFpEF trial (SUMMIT) showed numerically higher mortality with tirzepatide. Clinically meaningful improvements in quality of life and functional capacity were consistently observed in dedicated HFpEF trials. The strongest evidence supports GLP-1 RAs in HFpEF with obesity, where dedicated trials demonstrated clear benefits in HF events and symptoms. Future dedicated HF mortality trials with adjudicated endpoints are needed to determine whether the pooled mortality signal reflects a true HF-specific benefit.

## Data Availability

All data, analysis code, and materials necessary to reproduce the findings are publicly available in the Zenodo repository (DOI: 10.5281/zenodo.19100020). This includes raw search exports, screening decisions, extracted outcomes, risk of bias assessments, R analysis scripts, and rendered manuscript files.

https://zenodo.org/records/19100020

## Data Availability

All data, analysis code, and materials necessary to reproduce the findings are publicly available in the Zenodo repository (DOI: 10.5281/zenodo.18580111) and on GitHub (https://github.com/rodrigoeac/sr-glp1ra-hf).

## References

1. Savarese G, Becher PM, Lund LH, et al. Global burden of heart failure: A comprehensive and updated review of epidemiology. Cardiovascular Research. 2023; 118(17) :3272–3287.

2. Heidenreich PA, Bozkurt B, Aguilar D, et al. 2022 AHA/ACC/HFSA guideline for the management of heart failure. Journal of the American College of Cardiology. 2022; 79(17) :e263–e421.

3. McDonagh TA, Metra M, Adamo M, et al. 2021 ESC guidelines for the diagnosis and treatment of acute and chronic heart failure. European Heart Journal. 2021; 42(36) :3599–3726.

4. Borlaug BA, Jensen MD, Kitzman DW, et al. Obesity and heart failure with preserved ejection fraction: New insights and pathophysiological targets. Cardiovascular Research. 2023; 118(18) :3434–3450.

5. Drucker DJ. Mechanisms of action and therapeutic application of glucagon-like peptide-1. Cell Metabolism. 2018; 27(4) :740–756.

6. Ussher JR, Bhatt DL, Bhatt DL, Drucker DJ. Cardiovascular actions of incretin-based therapies. Circulation Research. 2023; 132(7) :888–904.

7. Marx N, Husain M, Lehrke M, Verma S, Sattar N. GLP-1 receptor agonists for the reduction of atherosclerotic cardiovascular risk in patients with type 2 diabetes. Circulation. 2022; 146(24) :1882–1894.

8. Marso SP, Daniels GH, Tanaka K, et al. Liraglutide and cardiovascular outcomes in type 2 diabetes. New England Journal of Medicine. 2016; 375(4) :311–322.

9. Holman RR, Bethel MA, Mentz RJ, et al. Effects of once-weekly exenatide on cardiovascular outcomes in type 2 diabetes. New England Journal of Medicine. 2017; 377(13) :1228–1239.

10. Hernandez AF, Green JB, Janmohamed S, et al. Albiglutide and cardiovascular outcomes in patients with type 2 diabetes and cardiovascular disease (Harmony Outcomes): A double-blind, randomised placebo-controlled trial. The Lancet. 2018; 392(10157) :1519–1529.

11. Gerstein HC, Colhoun HM, Dagenais GR, et al. Dulaglutide and cardiovascular outcomes in type 2 diabetes (REWIND): A double-blind, randomised placebo-controlled trial. The Lancet. 2019; 394(10193) :121–130.

12. Lincoff AM, Brown-Frandsen K, Colhoun HM, et al. Semaglutide and cardiovascular outcomes in obesity without diabetes. New England Journal of Medicine. 2023; 389(24) :2221–2232.

13. Kosiborod MN, Abildstrøm SZ, Borlaug BA, et al. Semaglutide in patients with heart failure with preserved ejection fraction and obesity. New England Journal of Medicine. 2023; 389(12) :1069–1084.

14. Kosiborod MN, Petrie MC, Borlaug BA, et al. Semaglutide in patients with obesity-related heart failure and type 2 diabetes. New England Journal of Medicine. 2024; 390(15) :1394–1407.

15. Packer M, Zile MR, Bolli A, et al. Tirzepatide for heart failure with preserved ejection fraction and obesity. New England Journal of Medicine. 2025; 392(5) :427–437.

16. Margulies KB, Hernandez AF, Redfield MM, et al. Effects of liraglutide on clinical stability among patients with advanced heart failure and reduced ejection fraction: A randomized clinical trial. JAMA. 2016; 316(5) :500–508.

17. Kong D, Li X, Zhang X, et al. Effects of glucagon-like peptide 1 receptor agonists on heart failure: A meta-analysis. Molecular Biology Reports. 2019; 46(2) :2397–2404.

18. Zhu C, Li J, et al. Glucagon-like peptide-1 receptor agonists and heart failure: An updated meta-analysis of randomized controlled trials. Clinical Cardiology. 2023; 46(12) :1484–1493.

19. Ji X, Wei Y, Qian J, et al. Effect of glucagon-like peptide-1 receptor agonists on heart failure in patients: A systematic review and meta-analysis. Diabetes Research and Clinical Practice. 2024; 208 :111097.

20. Page MJ, McKenzie JE, Bossuyt PM, et al. The PRISMA 2020 statement: An updated guideline for reporting systematic reviews. BMJ. 2021; 372 :n71.

21. Sterne JAC, Savović J, Page MJ, et al. RoB 2: A revised tool for assessing risk of bias in randomised trials. BMJ. 2019; 366 :l4898.

22. Hartung J, Knapp G. A refined method for the meta-analysis of controlled clinical trials with binary outcome. Statistics in Medicine. 2001; 20(24) :3875–3889.

23. Veroniki AA, Jackson D, Viechtbauer W, et al. Methods to estimate the between-study variance and its uncertainty in meta-analysis. Research Synthesis Methods. 2016; 7(1) :55–79.

24. DerSimonian R, Laird N. Meta-analysis in clinical trials. Controlled Clinical Trials. 1986; 7(3) :177–188.

25. Higgins JPT, Thompson SG, Deeks JJ, Altman DG. Measuring inconsistency in meta-analyses. BMJ. 2003; 327(7414) :557–560.

26. Riley RD, Higgins JPT, Deeks JJ. Interpretation of random effects meta-analyses. BMJ. 2011; 342 :d549.

27. Egger M, Davey Smith G, Schneider M, Minder C. Bias in meta-analysis detected by a simple, graphical test. BMJ. 1997; 315(7109) :629–634.

28. Duval S, Tweedie R. Trim and fill: A simple funnel-plot-based method of testing and adjusting for publication bias in meta-analysis. Biometrics. 2000; 56(2) :455–463.

29. Guyatt GH, Oxman AD, Vist GE, et al. GRADE: An emerging consensus on rating quality of evidence and strength of recommendations. BMJ. 2008; 336(7650) :924–926.

30. Balduzzi S, Rücker G, Schwarzer G. How to perform a meta-analysis with R: A practical tutorial. Evidence-Based Mental Health. 2019; 22(4) :153–160.

31. Viechtbauer W. Conducting meta-analyses in R with the metafor package. Journal of Statistical Software. 2010; 36(3) :1–48.

32. Ferreira JP, Saraiva F, Sharma A, et al. Albiglutide in patients with type 2 diabetes and heart failure: Harmony Outcomes trial post-hoc analysis. European Journal of Heart Failure. 2022; 24(7) :1248–1257.

33. Pfeffer MA, Claggett B, Diaz R, et al. Lixisenatide in patients with type 2 diabetes and acute coronary syndrome. New England Journal of Medicine. 2015; 373(23) :2247–2257.

34. Marso SP, Bain SC, Consoli A, et al. Semaglutide and cardiovascular outcomes in patients with type 2 diabetes. New England Journal of Medicine. 2016; 375(19) :1834–1844.

35. Husain M, Birkenfeld AL, Donsmark M, et al. Oral semaglutide and cardiovascular outcomes in patients with type 2 diabetes. New England Journal of Medicine. 2019; 381(9) :841–851.

36. McDonagh TA, Metra M, Adamo M, et al. 2023 focused update of the 2021 ESC guidelines for the diagnosis and treatment of acute and chronic heart failure. European Heart Journal. 2023; 44(37) :3627–3639.

37. McMurray JJV, Solomon SD, Inzucchi SE, et al. Dapagliflozin in patients with heart failure and reduced ejection fraction. New England Journal of Medicine. 2019; 381(21) :1995–2008.

38. Packer M, Anker SD, Butler J, et al. Cardiovascular and renal outcomes with empagliflozin in heart failure. New England Journal of Medicine. 2020; 383(15) :1413–1424.

39. Anker SD, Butler J, Filippatos G, et al. Empagliflozin in heart failure with a preserved ejection fraction. New England Journal of Medicine. 2021; 385(16) :1451–1461.

40. Solomon SD, McMurray JJV, Claggett B, et al. Dapagliflozin in heart failure with mildly reduced or preserved ejection fraction. New England Journal of Medicine. 2022; 387(12) :1089–1098.

