## supplement for "Glucagon-Like Peptide-1 Receptor Agonists Across the Heart Failure Spectrum: A Systematic Review and Meta-Analysis"

#### eTable 1. Search Strategies

##### PubMed (executed February 3, 2026; 697 records)

```
("glucagon-like peptide-1"[MeSH Terms] OR "glucagon-like peptide 1"[tiab]
 OR "GLP-1"[tiab] OR "GLP1"[tiab] OR "liraglutide"[tiab]
 OR "semaglutide"[tiab] OR "dulaglutide"[tiab] OR "exenatide"[tiab]
 OR "tirzepatide"[tiab] OR "lixisenatide"[tiab] OR "albiglutide"[tiab]
 OR "GLP-1 receptor agonist*"[tiab] OR "GLP-1RA"[tiab]
 OR "incretin mimetic*"[tiab])
AND
("heart failure"[MeSH Terms] OR "heart failure"[tiab]
 OR "cardiac failure"[tiab] OR "HFrEF"[tiab] OR "HFpEF"[tiab]
 OR "HFmrEF"[tiab] OR "preserved ejection fraction"[tiab]
 OR "reduced ejection fraction"[tiab] OR "systolic dysfunction"[tiab]
 OR "diastolic dysfunction"[tiab] OR "ventricular dysfunction"[tiab])
AND
("randomized controlled trial"[pt] OR "controlled clinical trial"[pt]
 OR "randomized"[tiab] OR "randomised"[tiab] OR "placebo"[tiab]
 OR "trial"[tiab])
```

No date or language restrictions. MeSH Terms used for primary concepts with free-text variants in title/abstract.

##### Cochrane CENTRAL (executed February 3, 2026; 356 records)

```
#1 [mh "Glucagon-Like Peptide 1"]
#2 (GLP-1 OR GLP1 OR semaglutide OR liraglutide OR tirzepatide
    OR dulaglutide OR exenatide):ti,ab,kw
```

```
#3 #1 OR #2

#4 [mh "Heart Failure"]

#5 ("heart failure" OR HFrEF OR HFpEF OR HFmrEF):ti,ab,kw

#6 #4 OR #5

#7 #3 AND #6
```

Filter: Cochrane CENTRAL (Trials) only.

#### ClinicalTrials.gov (executed February 3, 2026; 34 records)

```
Condition: Heart Failure

Intervention: GLP-1 OR semaglutide OR liraglutide OR tirzepatide

              OR dulaglutide OR exenatide

Study Type: Interventional (Clinical Trial)
```

#### eTable 2. Risk of Bias 2 Detailed Domain Assessments

*eTable 2. Risk of Bias 2 Detailed Domain Assessments*

| Study | D1: Randomisation | D2: Deviations | D3: Missing data | D4: Measurement | D5: Selection | Overall |
|---|---|---|---|---|---|---|
| SUMMIT | Low | Low | Low | Low | Low | Low |
| STEP-HFpEF | Low | Low | Low | Low | Low | Low |
| STEP-HFpEF-DM | Low | Low | Low | Low | Low | Low |
| FIGHT | Low | Low | Some concerns | Low | Low | Some concerns |
| LIVE | Low | Low | Low | Low | Low | Low |
| Albiglutide HFrEF | Some concerns | Low | Some concerns | Low | Low | Some concerns |
| SELECT (HF subgroup) | Low | Low | Low | Low | Some concerns | Some concerns |
| SOUL (HF analysis) | Low | Low | Low | Low | Some concerns | Some concerns |
| FLOW (HF subgroup) | Low | Low | Low | Low | Some concerns | Some concerns |
| EXSCEL (by EF) | Low | Low | Some concerns | Low | Some concerns | Some concerns |
| Harmony Outcomes (HF post-hoc) | Low | Low | Some concerns | Low | Some concerns | Some concerns |
| LEADER (HF subgroup) | Low | Low | Low | Low | Some concerns | Some concerns |
| REWIND (HF post-hoc) | Low | Low | Some concerns | Low | Some concerns | Some concerns |
| EXSCEL (by HF status) | Low | Low | Some concerns | Low | Some concerns | Some concerns |

D1: Randomisation process; D2: Deviations from intended interventions; D3: Missing outcome data; D4: Measurement of the outcome; D5: Selection of the reported result. For CVOT subgroup analyses, D5 was rated “Some concerns” because HF outcomes were typically secondary or exploratory endpoints.

#### eTable 3. GRADE Evidence Profile

*eTable 3. GRADE Evidence Profile*

| Outcome | N studies (participants) | Risk of Bias | Inconsistency | Indirectness | Imprecision | Publication Bias | Certainty |
|---|---|---|---|---|---|---|---|
| CV death + HHF | 8 (13143) | Not serious | Moderate ($I^2$=47%) | Serious[1] | Serious[2] | Undetected | Low |
| All-cause mortality | 7 (10137) | Not serious | Not serious ($I^2$=0%) | Serious[1] | Serious (incoherence)[3] | Undetected | Low |
| MACE | 4 | Not serious | Not serious ($I^2$=0%) | Serious[1] | Not serious | Undetected | Moderate |
| HF hospitalization | 5 | Not serious | Moderate ($I^2$=59%) | Serious[1] | Serious[2] | Undetected | Low |
| KCCQ-CSS | 3 | Not serious | Not serious ($I^2$=0%) | Not serious | Not serious | Undetected | Moderate |
| 6MWD | 4 | Not serious | Not serious ($I^2$=0%) | Not serious | Not serious | Undetected | Moderate |
| SAE (dedicated HF trials) | 4 (2176) | Not serious | Serious ($I^2$=78%) | Not serious | Serious[2] | Undetected | Low |

Footnotes: [1]Downgraded for indirectness: 8/14 studies are CVOT subgroup analyses where HF was not the primary endpoint. [2]Downgraded for imprecision: 95% CI crosses the null.

### eTable 4. Sensitivity Analysis Results (Primary Composite)

*eTable 4. Sensitivity Analyses for the Primary Composite Outcome*

| Analysis | k | HR | 95% CI | p | $I^2$ (%) |
|---|---|---|---|---|---|
| Primary (all studies) | 8 | 0.86 | [0.73, 1.01] | 0.0672 | 47.3 |
| Fixed-effect | 8 | 0.85 | [0.78, 0.94] | 0.0006 | 47.3 |
| Excl. Tirzepatide | 7 | 0.88 | [0.74, 1.04] | 0.1061 | 45.2 |
| Excl. FIGHT | 7 | 0.83 | [0.73, 0.94] | 0.0111 | 16.2 |
| Dedicated HF trials only | 2 | 0.91 | [0.01, 99.93] | 0.8359 | 86.0 |
| Excl. CVOT post-hoc subgroups | 2 | 0.91 | [0.01, 99.93] | 0.8359 | 86.0 |

### eFigure 1. Risk of Bias 2 Traffic Light Plot

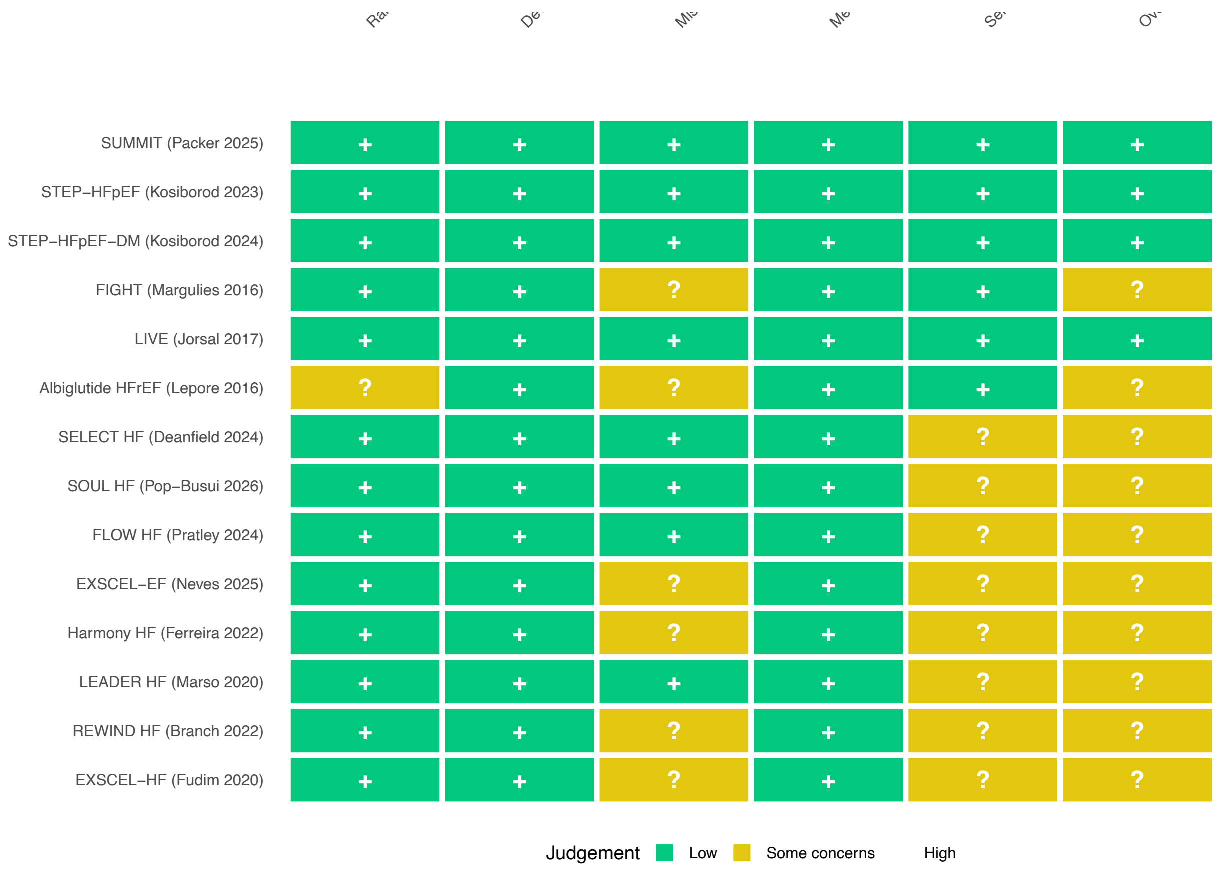


*eFigure 1. Risk of Bias 2 Traffic Light Plot*

#### eFigure 2. Risk of Bias 2 Summary Plot

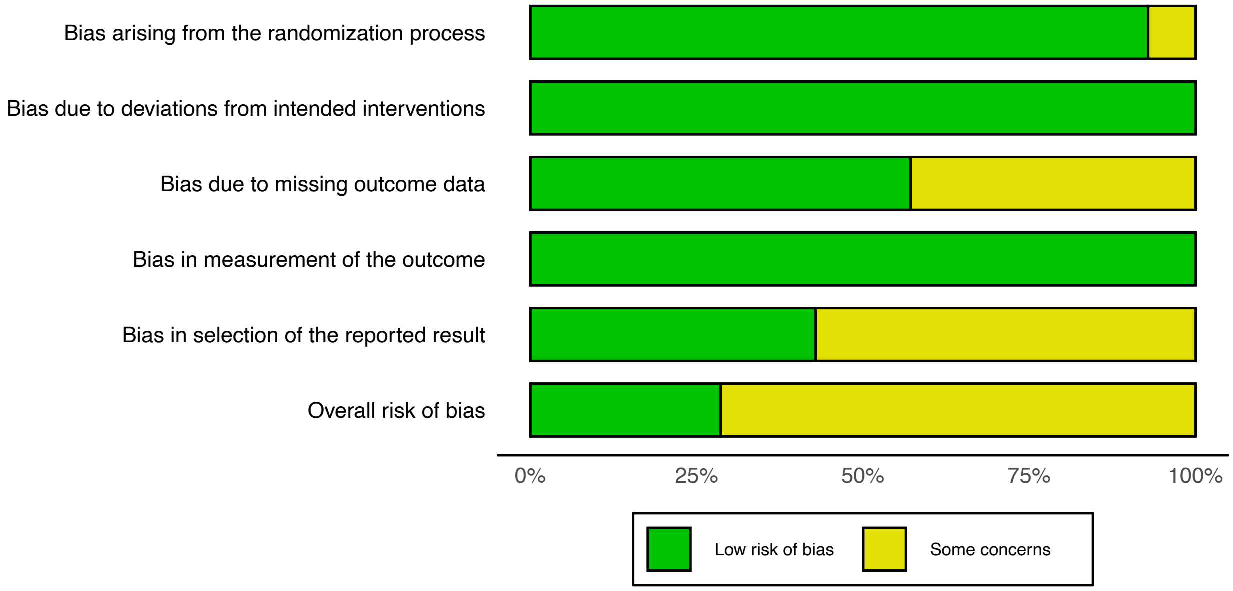


*eFigure 2. Risk of Bias 2 Summary Plot*

### eFigure 3. Forest Plot: Cardiovascular Death

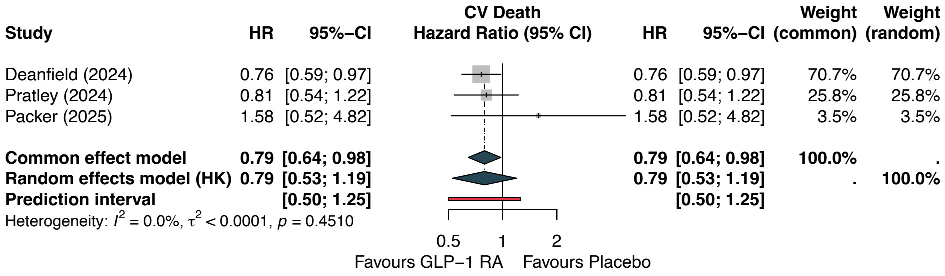


*eFigure 3. Forest plot of cardiovascular death.*

### eFigure 4. Forest Plot: Weight Change

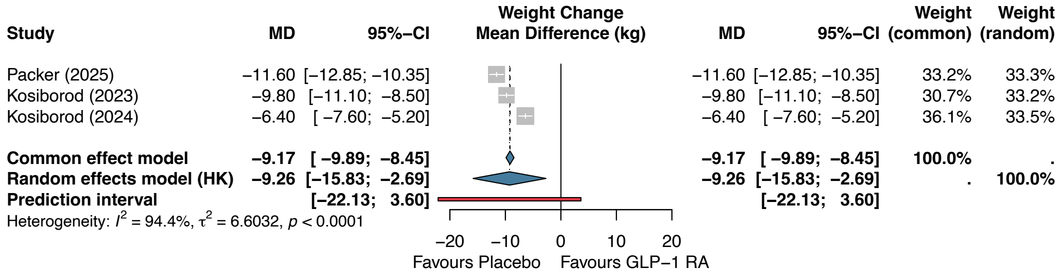


*eFigure 4. Forest plot of weight change.*

### eFigure 5. Forest Plot: LVEF Change

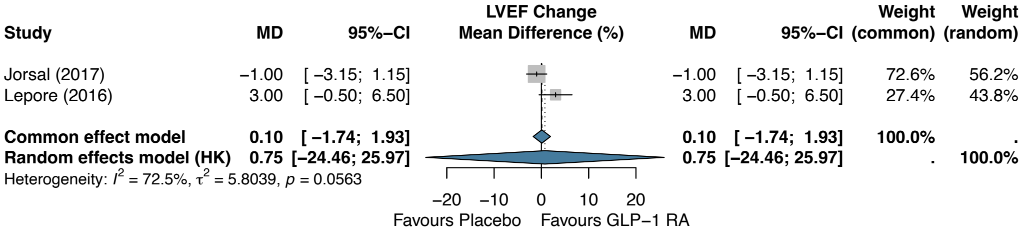


*eFigure 5. Forest plot of LVEF change.*

### eFigure 6. Forest Plot: Discontinuation Due to Adverse Events

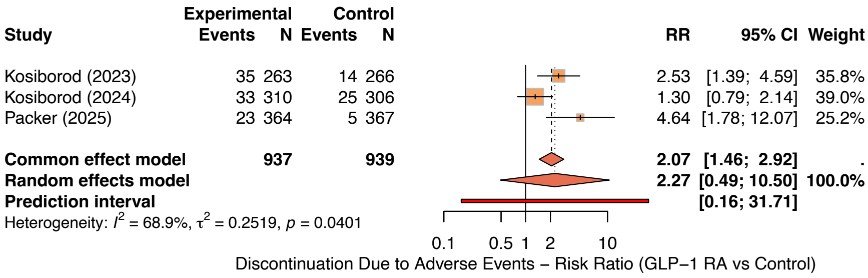


*eFigure 6. Forest plot of treatment discontinuation due to adverse events.*

### eFigure 7. Funnel Plot: Primary Composite Outcome

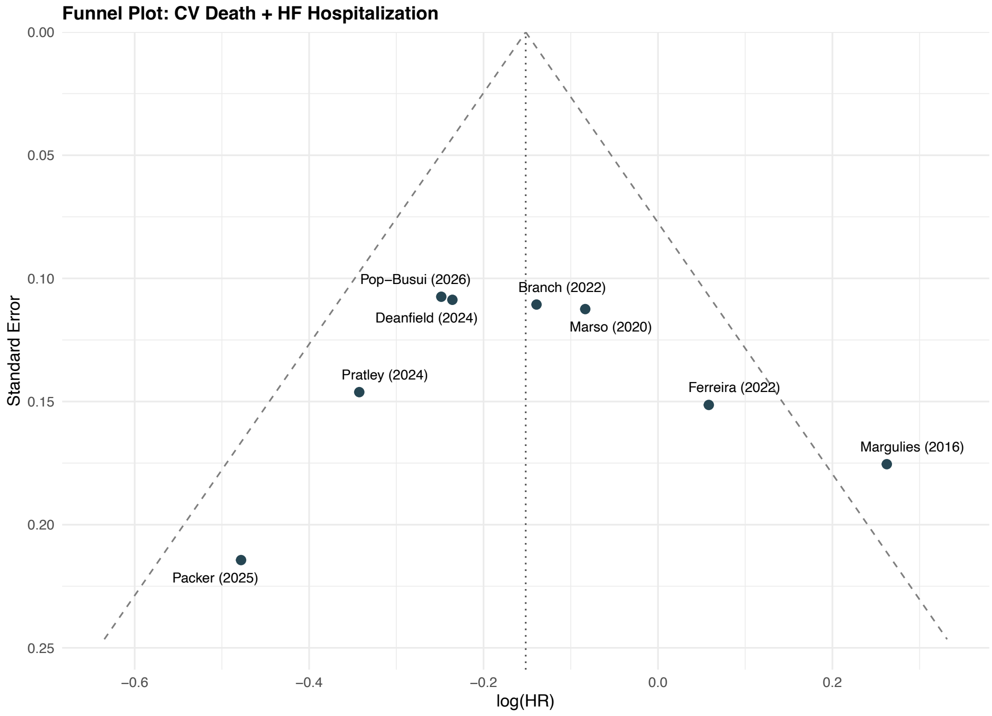


*eFigure 7. Funnel plot for the primary composite outcome.*

### eFigure 8. Funnel Plot with Trim-and-Fill

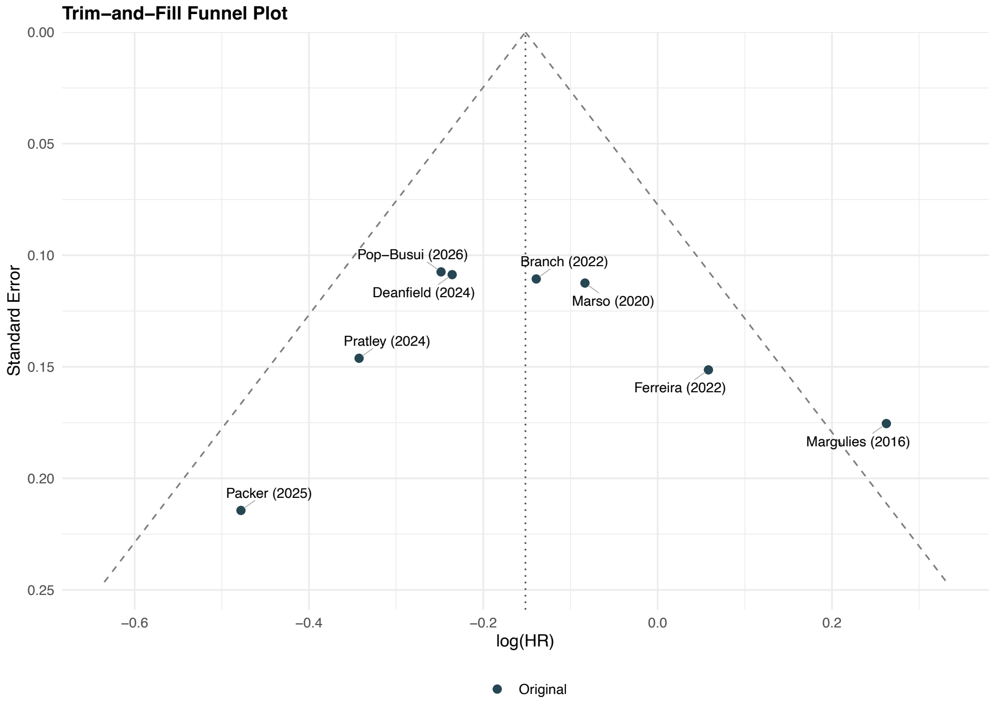


*eFigure 8. Funnel plot with trim-and-fill analysis.*

### eFigure 9. Leave-One-Out Sensitivity Analysis

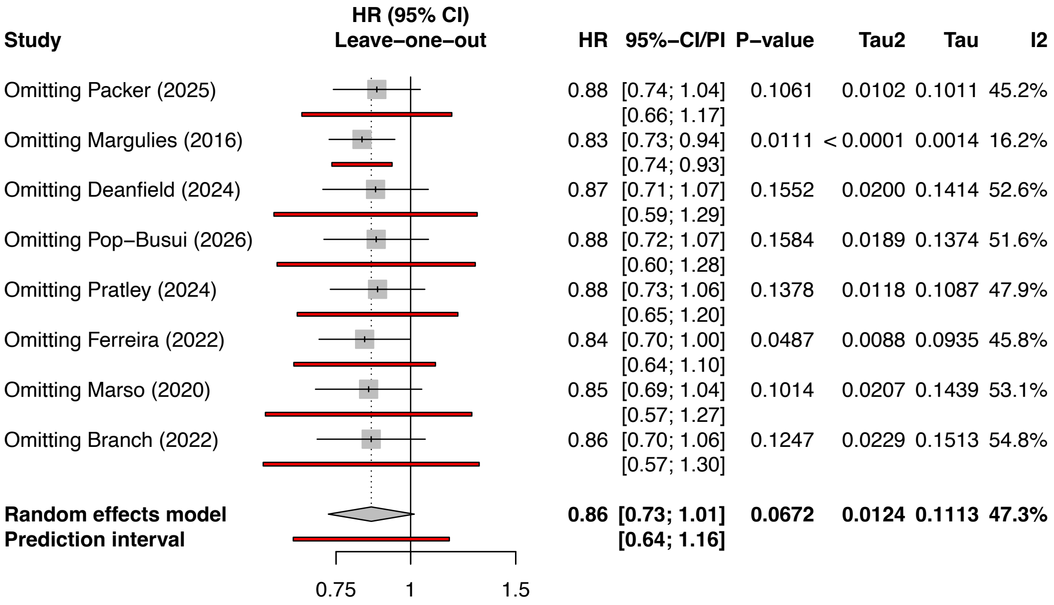


*eFigure 9. Leave-one-out sensitivity analysis for the primary composite outcome.*

### eFigure 10. Subgroup Analysis: HF Phenotype

| Study | logHR | SE(logHR) | HR (95% CI) | HR | 95%–CI | Weight (common) | Weight (random) |
|---|---|---|---|---|---|---|---|
| **hf_phenotype = HFpEF** | | | | | | | |
| Packer (2025) | −0.4780 | 0.2144 | | 0.62 | [0.41; 0.94] | 4.6% | 6.5% |
| **hf_phenotype = HFrEF** | | | | | | | |
| Margulies (2016) | 0.2624 | 0.1754 | | 1.30 | [0.92; 1.83] | 6.8% | 8.8% |
| **hf_phenotype = Mixed** | | | | | | | |
| Pratley (2024) | −0.3425 | 0.1462 | | 0.71 | [0.53; 0.95] | 9.8% | 11.3% |
| Pop−Busui (2026) | −0.2485 | 0.1075 | | 0.78 | [0.63; 0.96] | 18.2% | 16.0% |
| Deanfield (2024) | −0.2357 | 0.1087 | | 0.79 | [0.64; 0.98] | 17.8% | 15.8% |
| Branch (2022) | −0.1393 | 0.1106 | | 0.87 | [0.70; 1.08] | 17.1% | 15.5% |
| Marso (2020) | −0.0834 | 0.1125 | | 0.92 | [0.74; 1.15] | 16.6% | 15.2% |
| Ferreira (2022) | 0.0583 | 0.1514 | | 1.06 | [0.79; 1.43] | 9.2% | 10.8% |
| **Common effect model** | | | | **0.84** | **[0.76; 0.93]** | **88.6%** | **.** |
| **Random effects model (HK)** | | | | **0.84** | **[0.74; 0.96]** | **.** | **84.6%** |
| Heterogeneity: $I^2$ = 4.4%, $\tau^2$ = < 0.0001, $p$ = 0.3882 | | | | | | | |
| **Common effect model** | | | | **0.85** | **[0.78; 0.94]** | **100.0%** | **.** |
| **Random effects model (HK)** | | | | **0.86** | **[0.73; 1.01]** | **.** | **100.0%** |

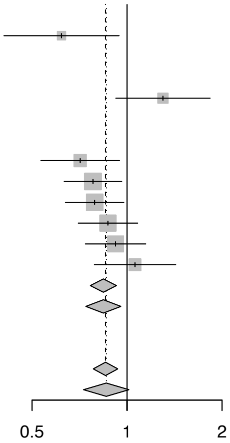


Heterogeneity: $I^2$ = 47.3%, $\tau^2$ = 0.0124, $p$ = 0.0653
Test for subgroup differences (common effect): $\chi^2_2$ = 8.06, df = 2 ($p$ = 0.0178)
Test for subgroup differences (random effects): $\chi^2_2$ = 8.06, df = 2 ($p$ = 0.0178)

*eFigure 10. Subgroup analysis by HF phenotype.*

### eFigure 11. Subgroup Analysis: Diabetes Status

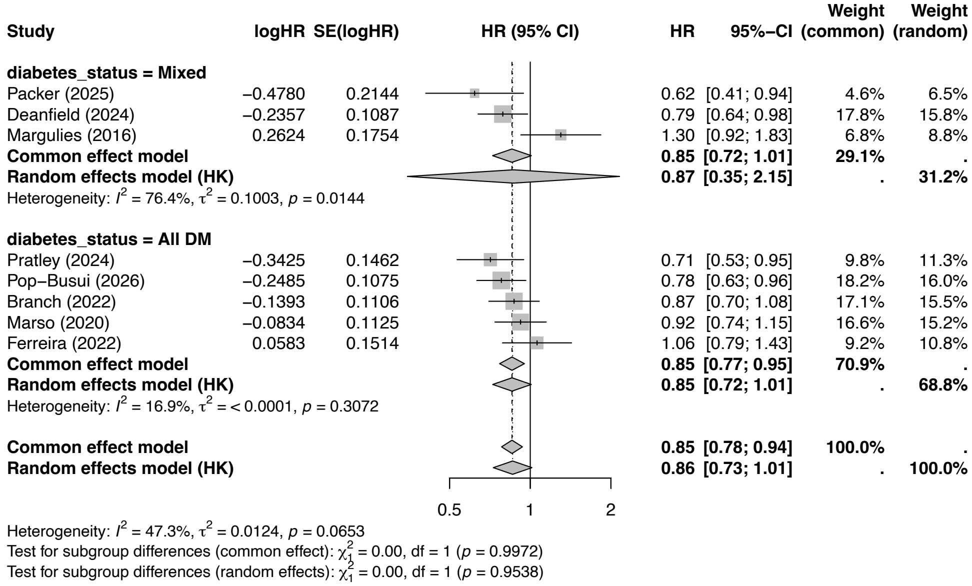


*eFigure 11. Subgroup analysis by diabetes status.*

### eFigure 12. Subgroup Analysis: GLP-1 RA Agent

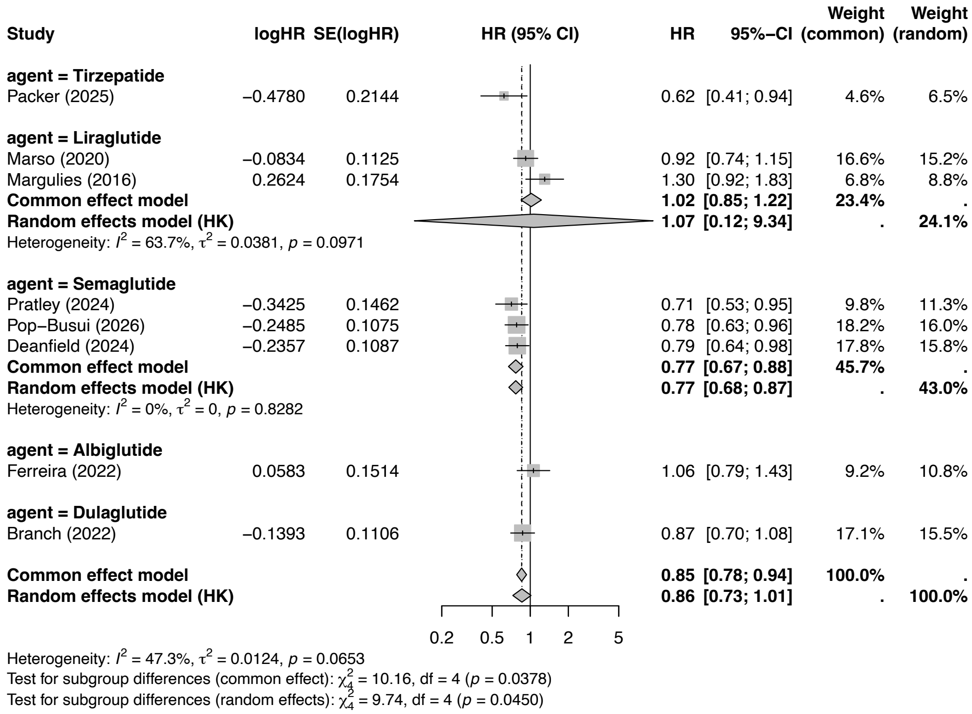


*eFigure 12. Subgroup analysis by GLP-1 RA agent.*

### eFigure 13. Subgroup Analysis: Study Type

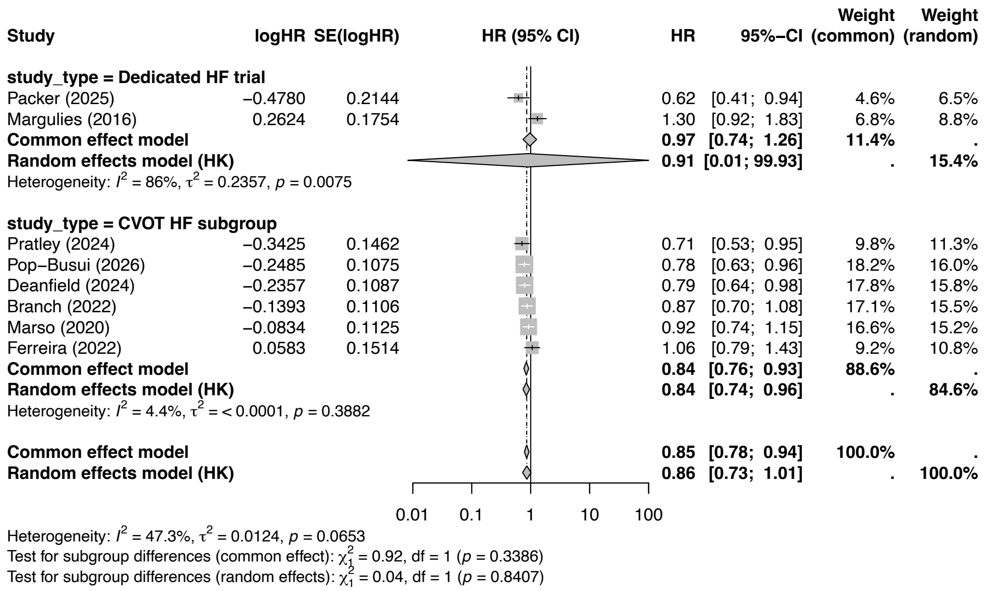


*eFigure 13. Subgroup analysis by study type.*

#### eTable 5. Composite Outcome Definitions by Study

| Study | Primary Composite Definition | Components | Notes |
|---|---|---|---|
| SUMMIT (Packer 2025) | CV death or worsening HF event | CV death, HF hospitalization, urgent HF visit, intensification of diuretics | Broader definition including outpatient worsening events |
| FIGHT (Margulies 2016) | Death or HF hospitalization | All-cause death, HF hospitalization | Includes all-cause (not CV-specific) death |
| SELECT HF (Deanfield 2024) | CV death or HF hospitalization | CV death, first HF hospitalization | Standard CVOT composite |
| SOUL HF (Pop-Busui 2026) | CV death or HF hospitalization | CV death, first HF hospitalization | Standard CVOT composite |

| Study | Primary Composite Definition | Components | Notes |
|---|---|---|---|
| FLOW HF (Pratley 2024) | CV death or HF hospitalization | CV death, first HF hospitalization | Standard CVOT composite |
| Harmony Outcomes HF (Ferreira 2022) | CV death or HF hospitalization | CV death, first HF hospitalization | Post-hoc subgroup |
| LEADER HF (Marso 2020) | CV death or HF hospitalization | CV death, first HF hospitalization | Pre-specified subgroup |
| REWIND HF (Branch 2022) | CV death or HF hospitalization | CV death, first HF hospitalization | Post-hoc subgroup |

Note: STEP-HFpEF, STEP-HFpEF-DM, LIVE, Albiglutide HFrEF, and EXSCEL subgroups did not report the primary composite outcome.

#### Amendments Log

##### Amendment 1: Search Strategy Re-execution (February 2026)

The initial database searches used simplified search terms that deviated from the full protocol-registered strategy. Upon recognition of this deviation, searches were re-executed using the complete protocol-compliant strategies (as documented in eTable 1). The screening process was reconstructed to ensure all records identified by the full strategy were assessed. No additional eligible studies were identified beyond those already captured by the initial searches.

##### Amendment 2: Screening Process (February 2026)

Initial title/abstract screening was performed using algorithmic filters (Python pipeline) rather than traditional independent dual-reviewer screening. All records passing initial filters (n=200) were subsequently assessed by both reviewers using the full PICO eligibility framework. This deviation from the standard dual-reviewer approach was documented and described transparently in the Methods section.

##### Amendment 3: EXSCEL Double-Counting (February 2026)

Two EXSCEL subgroup analyses (Neves by LVEF, Fudim by HF status) were included from the same parent trial. To avoid statistical dependence violations for secondary outcomes (ACM, MACE, HHF), the primary analysis uses only Neves et al. (by LVEF stratification, which more closely aligns with the PICO framework). Sensitivity analyses substituting Fudim et al. or excluding both were performed.

### eFigure 14. Summary of Binary Outcomes

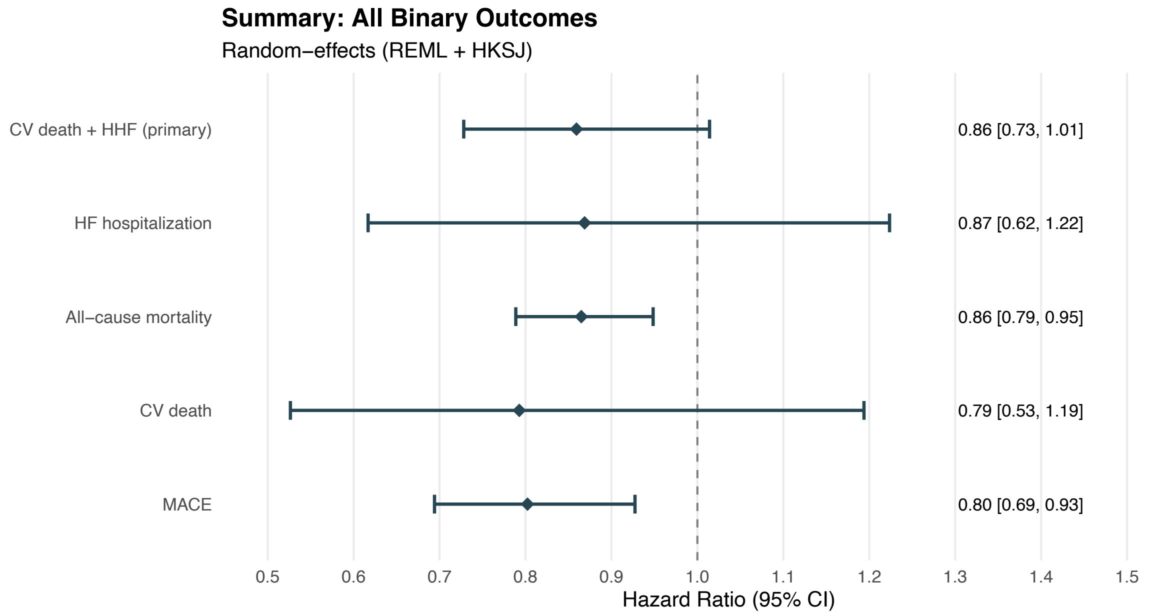


*eFigure 14. Summary forest plot of all binary outcomes.*

### eFigure 15. Summary of Continuous Outcomes

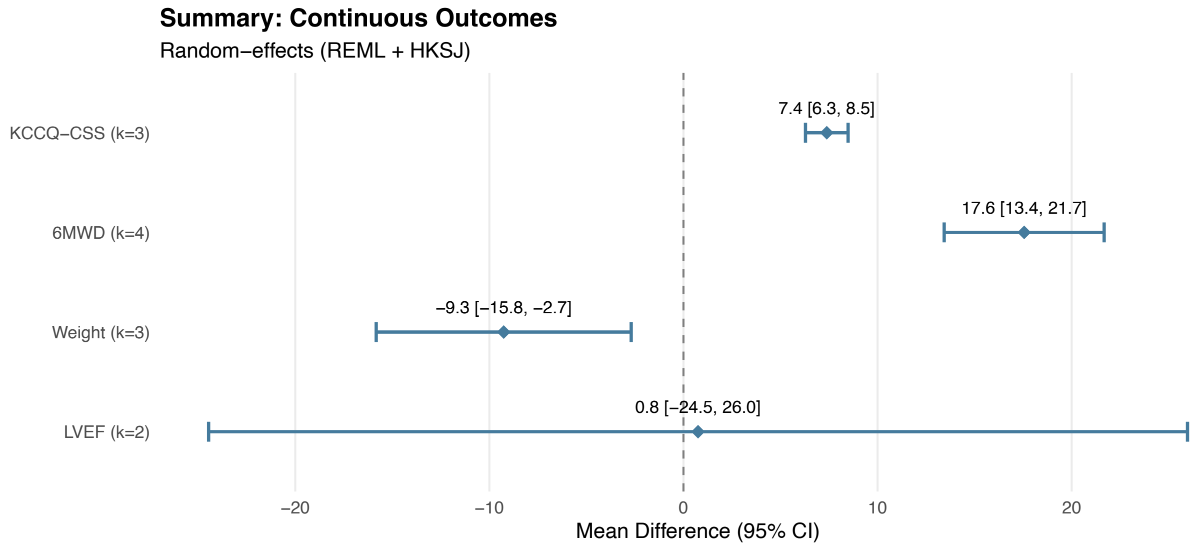


*eFigure 15. Summary forest plot of all continuous outcomes.*
